# Evaluation of Potential Adverse Events Following COVID-19 mRNA Vaccination Among Adults Aged 65 Years and Older: A Self-Controlled Study in the U.S

**DOI:** 10.1101/2023.01.19.23284803

**Authors:** Azadeh Shoaibi, Patricia C. Lloyd, Hui-Lee Wong, Tainya C. Clarke, Yoganand Chillarige, Rose Do, Mao Hu, Yixin Jiao, Andrew Kwist, Arnstein Lindaas, Rowan McEvoy, Michelle Ondari, Shruti Parulekar, Xiangyu Shi, Jing Wang, Yun Lu, Joyce Obidi, Cindy K. Zhou, Jeffrey A. Kelman, Richard A. Forshee, Steven A. Anderson

## Abstract

**Background:** Our near-real-time safety monitoring of 16 adverse events (AEs) following COVID-19 mRNA vaccination identified potential elevations in risk for six AEs following primary series and monovalent booster dose administration. The crude association with AEs does not imply causality. Accordingly, we conducted robust evaluations of the potential associations.

**Methods:** We conducted self-controlled case series studies of COVID-19 mRNA vaccines (BNT162b2 and mRNA-1273) in U.S. Medicare beneficiaries aged 65 years and older. Adjusted incidence rate ratio (IRRs) and 95% confidence intervals (CIs) were estimated following primary series doses for acute myocardial infarction (AMI), pulmonary embolism (PE), immune thrombocytopenia (ITP), disseminated intravascular coagulation (DIC); and following booster doses for AMI, PE, ITP, Bell ‘s Palsy (BP) and Myocarditis/Pericarditis (Myo/Peri).

**Results:** Among 3,360,981 individuals who received 6,388,542 primary series doses and 6,156,100 individuals with monovalent booster doses of either BNT162b2 or mRNA-1273, AE counts were: AMI (3,653 primary series, 16,042 booster), inpatient PE (2,470 primary, 5,085 booster), ITP (1,085 primary, 88 booster), DIC (254 primary), BP (3,268 booster), and Myo/Peri (1,295 booster). The IRR for inpatient PE cases following BNT162b2 primary series and booster was 1.19 (95% CI: 1.03 to 1.38) and 0.86 (95% CI: 0.78 to 0.95), respectively; and for mRNA-1273 primary series and booster, 1.15 (95% CI: 0.94 to 1.41) and 0.87 (95% CI: 0.79 to 0.96), respectively. The IRR for BP following BNT162b2 and mRNA-1273 booster was 1.17 (95% CI: 1.06 to 1.29) and 1.16 (95% CI: 1.05 to 1.29), respectively.

**Conclusion:** In these two studies of the U.S. elderly we did not find an increased risk for AMI, ITP, DIC, and Myo/Peri; the results were not consistent for PE; and there was a small elevated risk of BP after exposure to COVID-19 mRNA vaccines. These results support the favorable safety profile of COVID-19 mRNA vaccines administered in the elderly.

**HIGHLIGHTS:** There was no increased risk for four of six outcomes following COVID-19 monovalent mRNA vaccines. There was a small elevated risk of Bell ‘s Palsy after exposure to COVID-19 monovalent mRNA vaccines. Risk of pulmonary embolism was not consistent after exposure to COVID-19 monovalent mRNA vaccines.

## 1 INTRODUCTION

The coronavirus disease 2019 (COVID-19) pandemic heavily impacted both the United States (U.S.) and global populations. Since the start of the pandemic there have been a total of about 100 million COVID-19 cases, nearly 1.1 million related deaths and 5.8 million related hospitalizations in the U.S.^1,2^ Of the observed COVID-19-related deaths, about 75 percent have occurred in individuals aged 65 and older.^3^ In response, the U.S. Food and Drug Administration (FDA) has authorized (under emergency use authorization) or approved COVID-19 vaccines for prevention of COVID-19 including primary series (doses 1 and 2) and additional booster doses.^4^ These include the Pfizer BioNTech vaccine available for persons 6 months and older (BNT162b2), Moderna vaccine available for persons 6 months and older (mRNA-1273) and the Novavax vaccine (NVX-CoV2373) available for persons 12 years and older.^5-7^ The monovalent vaccines available in the U.S. are based on the original strain of the SARS-CoV-2 virus.^8^

The FDA conducts active post-market surveillance to monitor the safety of COVID-19 vaccines (i) primary series and (ii) booster doses in all ages including those 65 years and older. Near real-time surveillance or rapid cycle analyses (RCA) is a sequential testing method to screen for an increased risk of adverse events (AEs) following vaccination as vaccination data accrue. Through this framework using the U.S. Centers for Medicare and Medicaid Services (CMS) Medicare database, the observed incidence rates of 16 pre-specified AEs were compared to historical AEs rates following administration of available COVID-19 mRNA vaccines during the study period.

This rapid screening method in persons 65 years and older identified statistically significant associations (signals) between the primary series BNT162b2 vaccine and acute myocardial infarction (AMI), pulmonary embolism (PE), disseminated intravascular coagulation (DIC), and immune thrombocytopenia (ITP).^9^ A signal detection study of COVID-19 monovalent booster dose vaccines (BNT162b2, mRNA-1273) detected signals for BNT162b2 COVID-19 vaccine and Bell ‘s Palsy (BP), as well as for mRNA-1273 COVID-19 vaccine and myocarditis/pericarditis (Myo/Peri). These six events may not be true safety concerns as the RCA cannot establish that the vaccines caused these AEs. While preliminary signal detection studies enable rapid safety screening, they can be subject to bias and confounding and must be further evaluated in signal evaluation studies that more rigorously adjust for various sources of confounding.

This manuscript summarizes results from two independent vaccine safety studies that used a self-controlled study design to account for time-invariant confounders. Specifically, we estimated the risk of AMI, PE, DIC and ITP following primary series vaccination with the COVID-19 mRNA vaccines, BNT162b2 and mRNA-1273, in a study referred to as the ‘primary series study ‘ as well as the risk of AMI, PE, ITP, BP, and Myo/Peri following COVID-19 mRNA monovalent booster vaccination in a study referred to as the ‘booster study ‘.

## 2 METHODS

### 2.1 Study Design, Data Sources and Study Period

We define primary series as doses 1 and 2 of a COVID-19 mRNA vaccine, and the booster dose as a subsequent (or third) monovalent dose following a COVID-19 mRNA vaccine primary series. In this study, COVID-19 bivalent mRNA vaccines were not evaluated. Third doses (or first monovalent booster doses) are hereafter referred to as ‘booster ‘ doses for clarity. We compared the incidence of AEs within periods of hypothesized excess risk due to vaccine administration (risk interval) with their incidence during a control interval for COVID-19 primary series and booster vaccines, in two separate studies, among Medicare Fee-For-Service (FFS) individuals aged 65 years and older. Self-controlled case series (SCCS) or self-controlled risk interval (SCRI) analyses were conducted to compare exposed and unexposed periods within the same individual, and thus inherently adjusted for sources of time-invariant confounding.^10^ The statistical analytical plan was pre-specified in the protocol and summarized in Table 1. Details of the study specifications are outlined in eTable 1.^11^

**Table 1.**
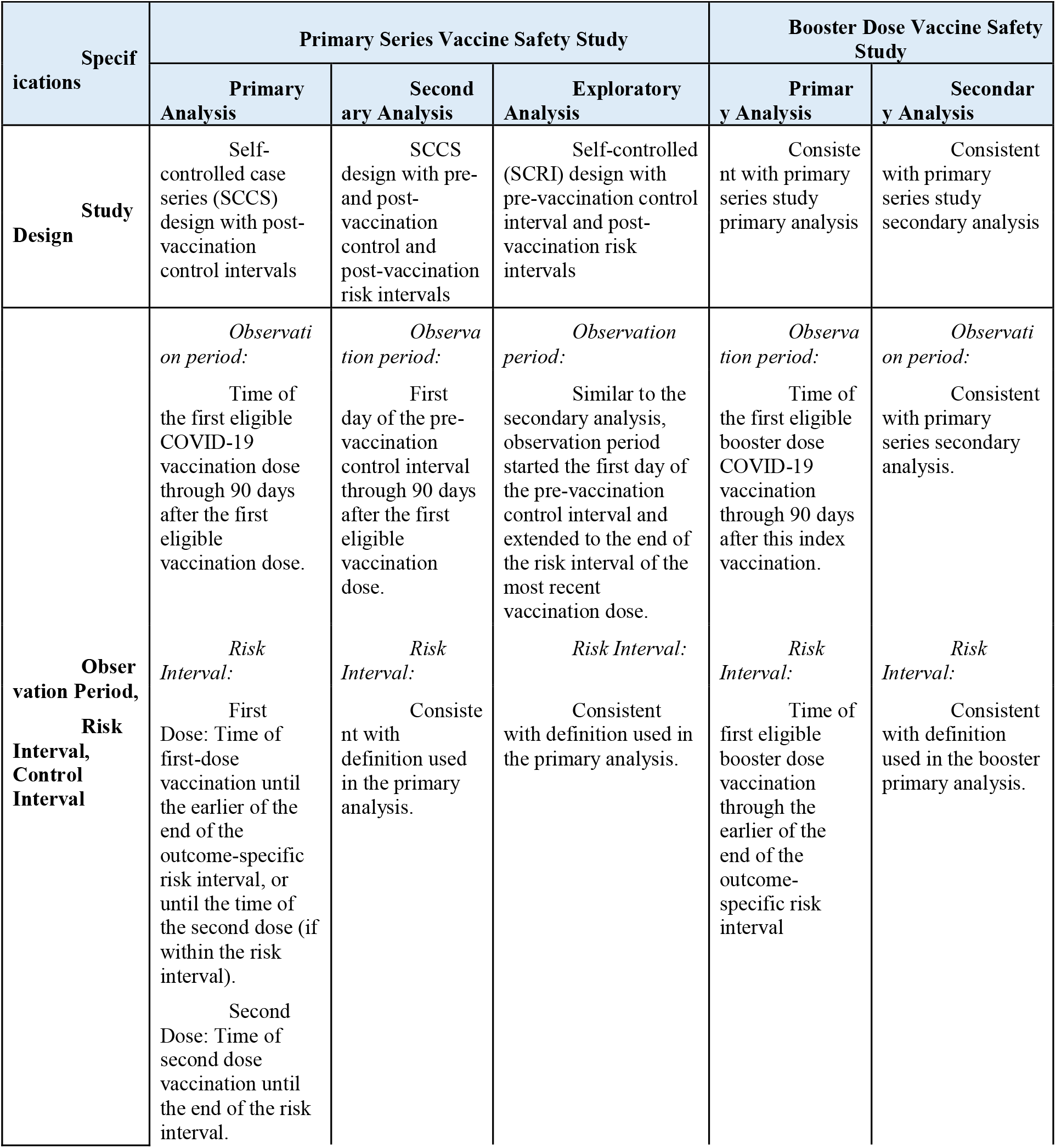
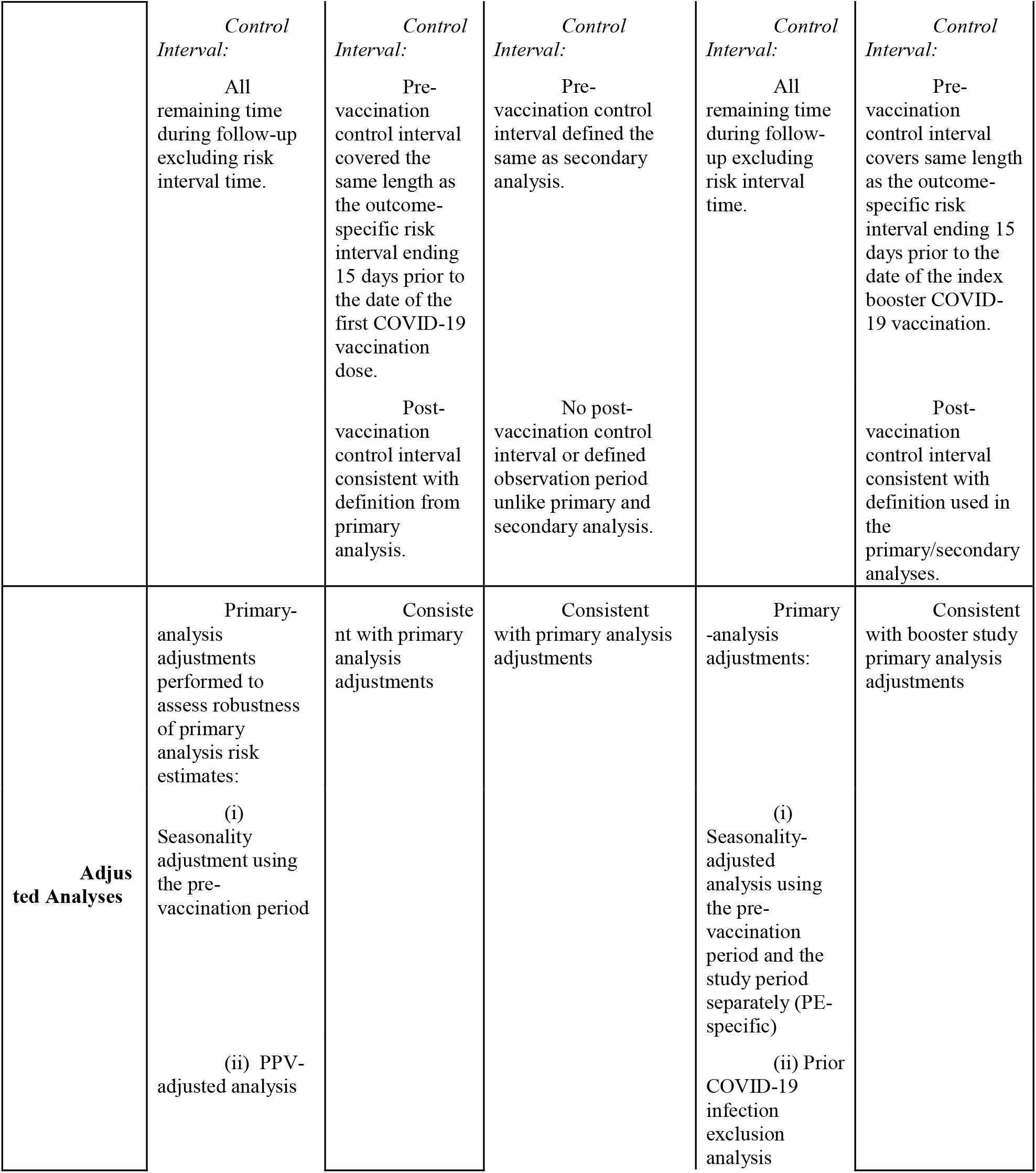
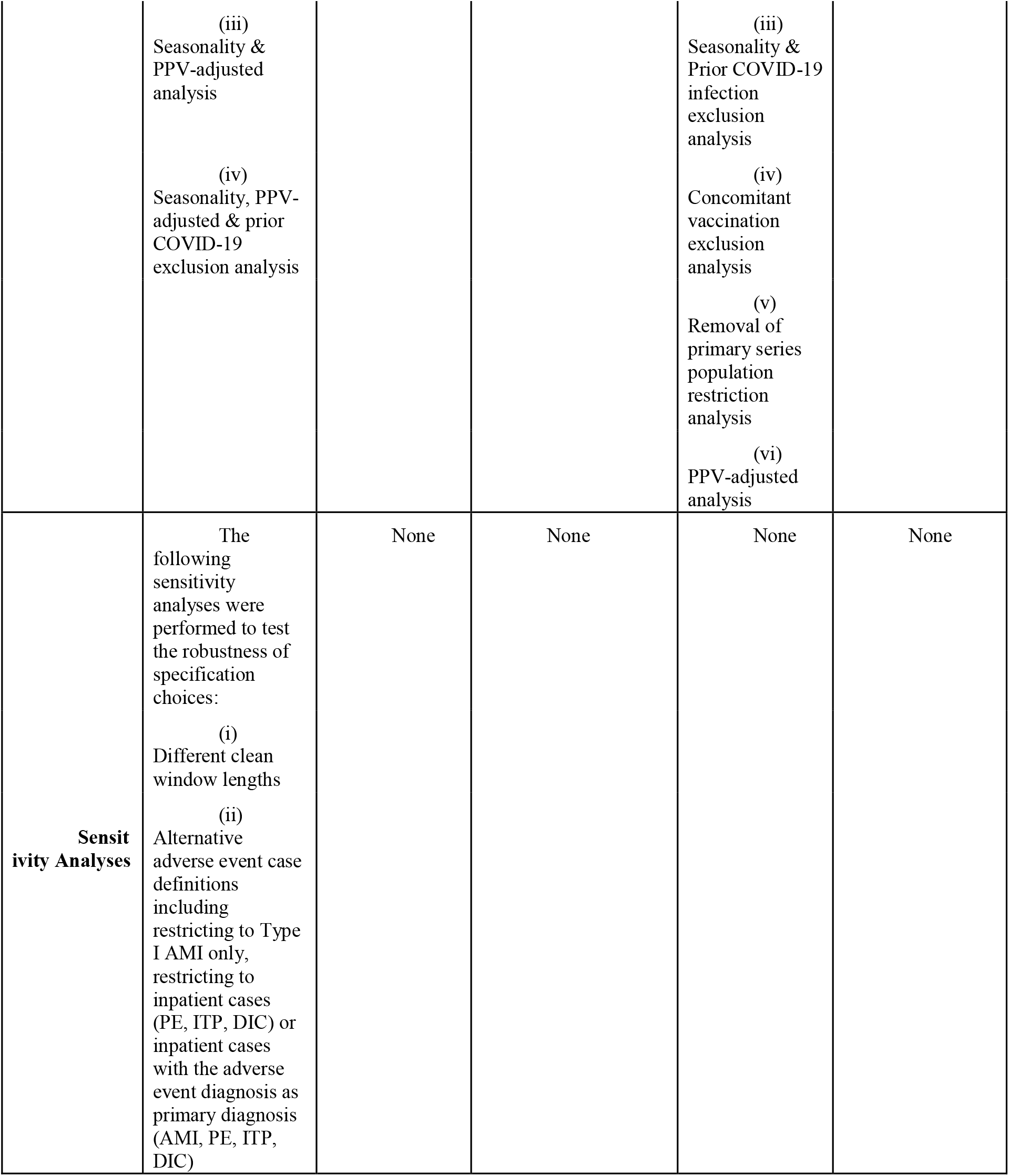
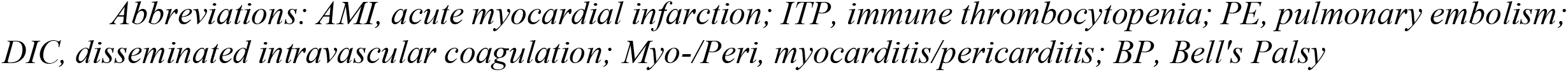
Summary of Observation Period and Analyses for Primary, Secondary, Exploratory and Subgroup Analyses and Associated Adjusted Analyses in the COVID-19 Primary Series and Booster Dose Vaccine Safety Studies.

| Specifications | Primary Series Vaccine Safety Study |  |  | Booster Dose Vaccine Safety Study |  |
| --- | --- | --- | --- | --- | --- |
|  | Primary Analysis | Secondary Analysis | Exploratory Analysis | Primary Analysis | Secondary Analysis |
| **Study Design** | Self-controlled case series (SCCS) design with post-vaccination control intervals | SCCS design with pre- and post-vaccination control and post-vaccination risk intervals | Self-controlled (SCRI) design with pre-vaccination control interval and post-vaccination risk intervals | Consistent with primary series study primary analysis | Consistent with primary series study secondary analysis |
| **Observation Period, Risk Interval, Control Interval** | *Observation period:*<br>Time of the first eligible COVID-19 vaccination dose through 90 days after the first eligible vaccination dose.<br>*Risk Interval:*<br>First Dose: Time of first-dose vaccination until the earlier of the end of the outcome-specific risk interval, or until the time of the second dose (if within the risk interval).<br>Second Dose: Time of second dose vaccination until the end of the risk interval. | *Observation period:*<br>First day of the pre-vaccination control interval through 90 days after the first eligible vaccination dose.<br>*Risk Interval:*<br>Consistent with definition used in the primary analysis. | *Observation period:*<br>Similar to the secondary analysis, observation period started the first day of the pre-vaccination control interval and extended to the end of the risk interval of the most recent vaccination dose.<br>*Risk Interval:*<br>Consistent with definition used in the primary analysis. | *Observation period:*<br>Time of the first eligible booster dose COVID-19 vaccination through 90 days after this index vaccination.<br>*Risk Interval:*<br>Time of first eligible booster dose vaccination through the earlier of the end of the outcome-specific risk interval | *Observation period:*<br>Consistent with primary series secondary analysis.<br>*Risk Interval:*<br>Consistent with definition used in the booster primary analysis. |
|  | *Control Interval:*<br>All remaining time during follow-up excluding risk interval time. | *Control Interval:*<br>Pre-vaccination control interval covered the same length as the outcome-specific risk interval ending 15 days prior to the date of the first COVID-19 vaccination dose.<br>Post-vaccination control interval consistent with definition from primary analysis. | *Control Interval:*<br>Pre-vaccination control interval defined the same as secondary analysis.<br>No post-vaccination control interval or defined observation period unlike primary and secondary analysis. | *Control Interval:*<br>All remaining time during follow-up excluding risk interval time. | *Control Interval:*<br>Pre-vaccination control interval covers same length as the outcome-specific risk interval ending 15 days prior to the date of the index booster COVID-19 vaccination.<br>Post-vaccination control interval consistent with definition used in the primary/secondary analyses. |
| **Adjusted Analyses** | Primary-analysis adjustments performed to assess robustness of primary analysis risk estimates:<br>(i) Seasonality adjustment using the pre-vaccination period<br>(ii) PPV-adjusted analysis | Consistent with primary analysis adjustments | Consistent with primary analysis adjustments | Primary-analysis adjustments:<br>(i) Seasonality-adjusted analysis using the pre-vaccination period and the study period separately (PE-specific)<br>(ii) Prior COVID-19 infection exclusion analysis | Consistent with booster study primary analysis adjustments |
|  | (iii)<br>Seasonality & PPV-adjusted analysis<br>(iv)<br>Seasonality, PPV-adjusted & prior COVID-19 exclusion analysis |  |  | (iii)<br>Seasonality & Prior COVID-19 infection exclusion analysis<br>(iv)<br>Concomitant vaccination exclusion analysis<br>(v)<br>Removal of primary series population restriction analysis<br>(vi)<br>PPV-adjusted analysis |  |
| **Sensitivity Analyses** | The following sensitivity analyses were performed to test the robustness of specification choices:<br>(i)<br>Different clean window lengths<br>(ii)<br>Alternative adverse event case definitions including restricting to Type I AMI only, restricting to inpatient cases (PE, ITP, DIC) or inpatient cases with the adverse event diagnosis as primary diagnosis (AMI, PE, ITP, DIC) | None | None | None | None |

*Abbreviations: AMI, acute myocardial infarction; ITP, immune thrombocytopenia; PE, pulmonary embolism; DIC, disseminated intravascular coagulation; Myo-/Peri, myocarditis/pericarditis; BP, Bell's Palsy*

We utilized longitudinal claims and enrollment data from the Medicare database to obtain information on beneficiaries ‘ demographics, enrollment, vaccination history, medical history, AE occurrence, and nursing home residence status.

The study start date for the primary series and booster studies aligned with the earliest Emergency Use Authorization (EUA) date for the respective vaccines. Study end dates for both sets of vaccines were independently specified as the dates when the study achieved sufficient power to detect a minimum pre-specified risk, and to ensure at least 90 percent data completeness. For the primary series study, the start date was December 11, 2020 and the event-specific study end dates were April 16, 2021 for AMI, April 30, 2021 for PE and DIC, and May 7, 2021 for ITP. The booster study start date was August 12, 2021, and the event-specific study end dates were April 30, 2022 for AMI, PE and ITP, May 7, 2022 for Myo/Peri, and May 14, 2022 for BP.

### 2.2 Study Population, Exposure, Adverse Events, and Follow-Up

The primary series and booster studies both included elderly Medicare FFS beneficiaries who received at least one of the specified COVID-19 mRNA vaccine doses and experienced an incident AE occurrence during the follow-up. The booster study excluded individuals without an observed primary series vaccination in alignment with EUA authorized uses.

Exposure and AE definitions are found in eTables 1 and 2, respectively. The six AEs in the study were identified from previously performed active monitoring surveillance analyses specific to the primary series and booster dose vaccines. The primary series study focused on the post-vaccination risk of AMI, DIC, and PE using a risk window of 1-28 days, and ITP using a risk window of 1-42 days post-vaccination. The booster study examined the risk of AMI and inpatient PE using a 1-28 day post-vaccination risk window, BP and ITP using a 1-42 day post-vaccination risk window, and Myo/Peri using a 1-21 day post-vaccination risk window. While consistent event definitions were used for AMI and PE in the two studies, the booster study used a more restrictive inpatient ITP event definition to improve the specificity of cases identified.

To ensure sufficient observation time, beneficiaries were required to accrue follow-up time during both the risk and control intervals, unless death occurred before the control window. Beneficiaries were followed until the earliest occurrence of observation period end, study period end, disenrollment, death, or a fourth vaccine dose. Continuous enrollment was required from the clean window prior to the occurrence of an AE through follow-up, and in the 365 days prior to the incident AE. To assess the effect of individual vaccine brands and prevent misclassification of vaccine doses, both studies excluded individuals with heterologous vaccination use, and vaccinations too close in proximity to previous vaccine doses.

### 2.3 Medical Record Review

To validate the claims-based AE definitions, medical record review (MRR) was conducted for cases identified from the primary series (AMI, PE (all-settings, IP-only), ITP (all settings), DIC) and booster studies (BP, ITP (IP-only, primary diagnosis) Myo/Peri). For each case definition, medical records were obtained and adjudicated from a random sample of cases identified in the study. Cases were then classified as true cases, non-cases, and indeterminate, using standard clinical definitions. For each AE definition, a positive predictive value (PPV) along with a corresponding 95% confidence interval (CI) was estimated.^12^ Table 3 presents classification decisions and PPV estimates by AE. These estimates were used to conduct a quantitative bias analysis (QBA) for each AE to assess the direction and magnitude of event misclassification.^13^

To facilitate timely analysis, PPV estimates from the primary series MRR were utilized in the booster study QBA for AMI and PE. Additional MRR from the booster study was initiated for BP, ITP (IP-only, primary diagnosis), Myo/Peri and is ongoing, and the results are not available for this manuscript. MRR of the ITP (all settings) definition was also performed in the primary series study.

### 2.4 Statistical Analysis

Descriptive statistics were calculated for categorical variables. The primary analysis used an SCCS study design with a post-vaccination control interval. In the primary analysis, follow-up included all time up to 90 days post-vaccination, with post-vaccination time in pre-specified risk windows considered exposed time and all remaining time considered control time. A conditional Poisson regression was used to estimate the incidence rate ratio (IRR) comparing rates in the risk and control intervals for each AE and the corresponding attributable risk (AR). The AR was calculated by the excess number of cases predicted from the regression model divided by the number of eligible vaccinations or person-time.^14^

The secondary analysis included both pre-vaccination and post-vaccination control intervals and was performed to evaluate the robustness of risk estimates from the primary analysis to variations in the period used to estimate baseline risk. Additional control windows from 15 to 43 days pre-vaccination were included.

Given the high fatality rate of certain events, an adjustment to address bias from event-dependent observation time was conducted.^15^ Adjustments to the primary and secondary analyses were performed to further investigate the impact of various potential sources of confounding: (i) a seasonality adjustment to account for potential time-varying confounding due to seasonal changes in incidence rates, (ii) an analysis using the PPV from MRR to conduct QBA to assess robustness of results to event misclassification, and (iii) an analysis excluding individuals with prior COVID-19 infection to account for the hypothesized association between the infection and AEs. ^16-23^

Additionally, there were analyses unique to each evaluation. The primary series study included (i) an exploratory analysis using only the pre-vaccination control interval to assess robustness to temporal variations in baseline risk, and (ii) adjusted analyses varying the definition of events, including care settings as well as risk and control intervals. The booster study included (i) an analysis removing the requirement that a primary series be observed given the limited observability of primary series vaccinations in the Medicare population, and (ii) a PE-specific post-hoc analysis adjusting for seasonality using pandemic rather than pre-pandemic incidence rates. All analyses were conducted using R 4.0.3 (R Foundation for Statistical Computing, Vienna, Austria), and SAS v. 9.4 (SAS Institute Inc., Cary, NC, United States). This surveillance activity was conducted as part of the FDA public health surveillance mandate.

## 3 RESULTS

### 3.1 Descriptive Results

We evaluated the risk of AMI, PE, DIC, and ITP in a study of the primary series doses of COVID-19 mRNA vaccines (primary series study) and evaluated the risk of AMI, PE, ITP, BP, and Myo/Peri in a separate monovalent booster dose study (booster study). Table 2 summarizes the baseline characteristics of vaccinated individuals who developed an AE in the follow-up windows for both studies. Among 3,360,981 individuals who received 6,388,542 primary series doses and 6,156,100 individuals with monovalent booster doses of either BNT162b2 or mRNA-1273, case counts were as follows: AMI (3,653 primary series, 16,042 booster), inpatient PE (2,470 primary series, 5,085 booster), ITP (1,085 primary series, 88 booster), DIC (254 primary series), BP (3,268 booster), and Myo/Peri (1,295 booster). In both studies, BNT162b2 recipients compared to mRNA-1273 recipients were generally younger, less likely to reside in a rural area, less likely to be dual-eligible for Medicare and Medicaid, and more likely to reside in a nursing home. The primary series study had a higher proportion of cases that were older, residing in a nursing home, and eligible for dual Medicare and Medicaid compared to the booster study (Table 2).

**Table 2.**
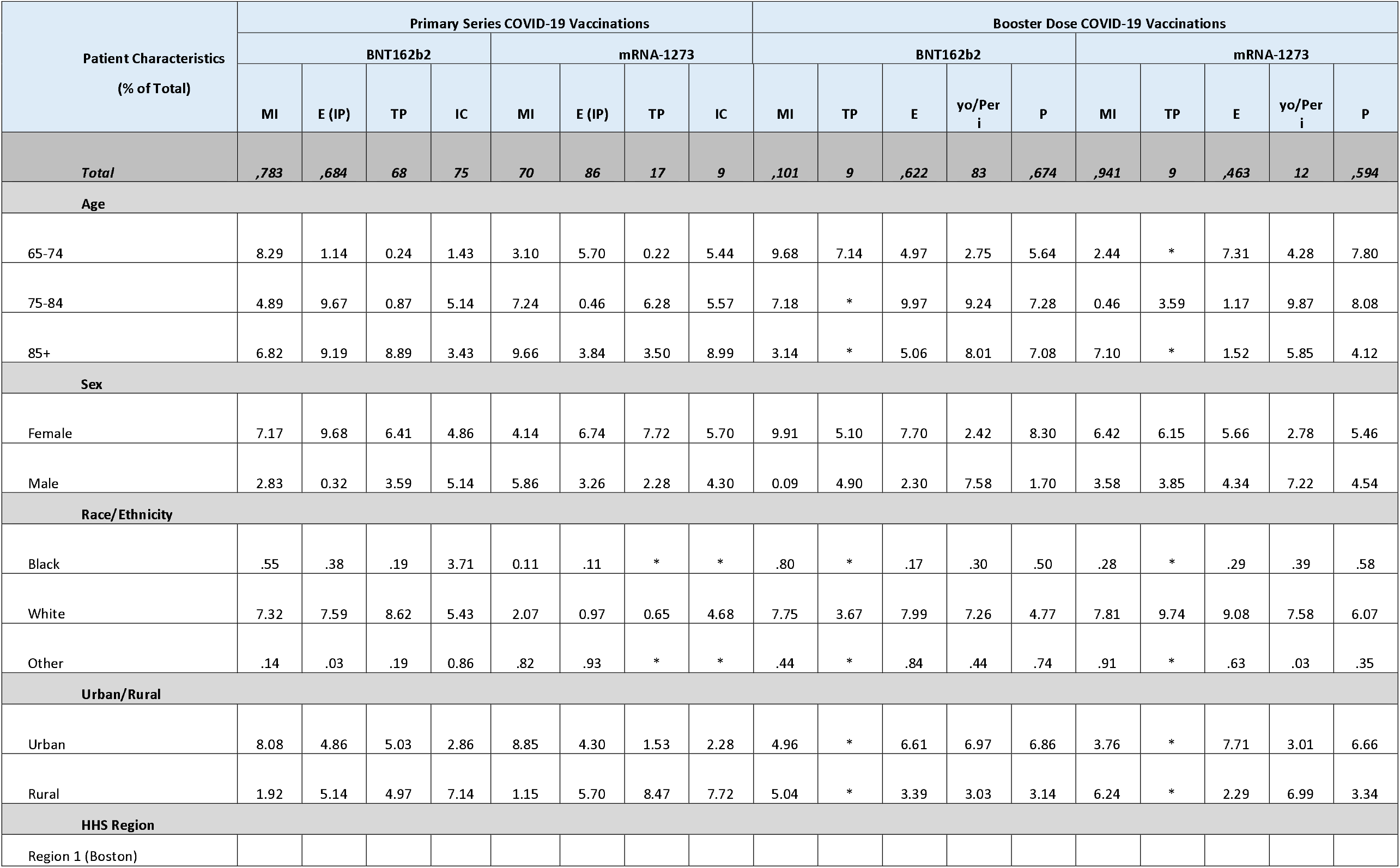
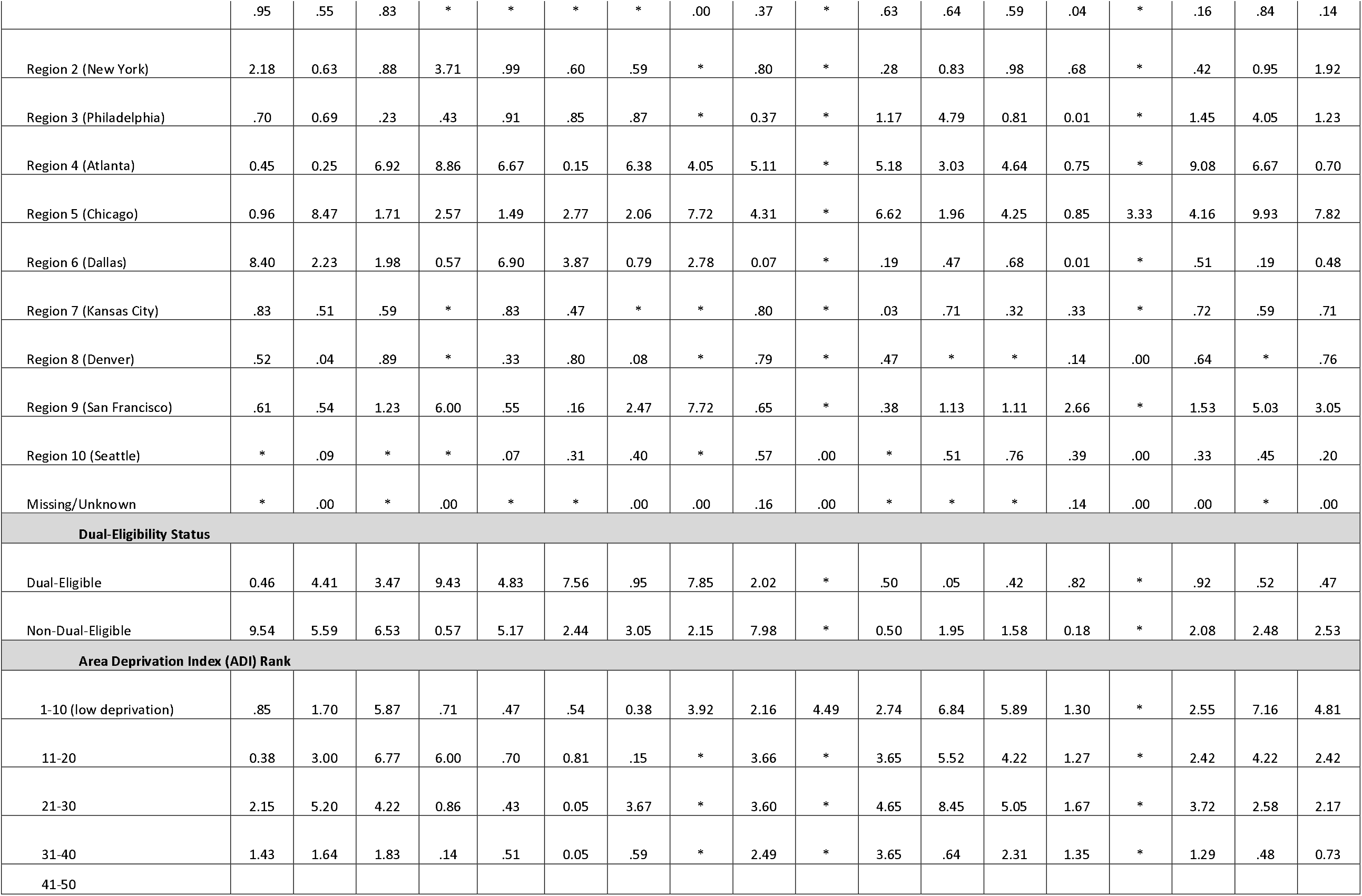
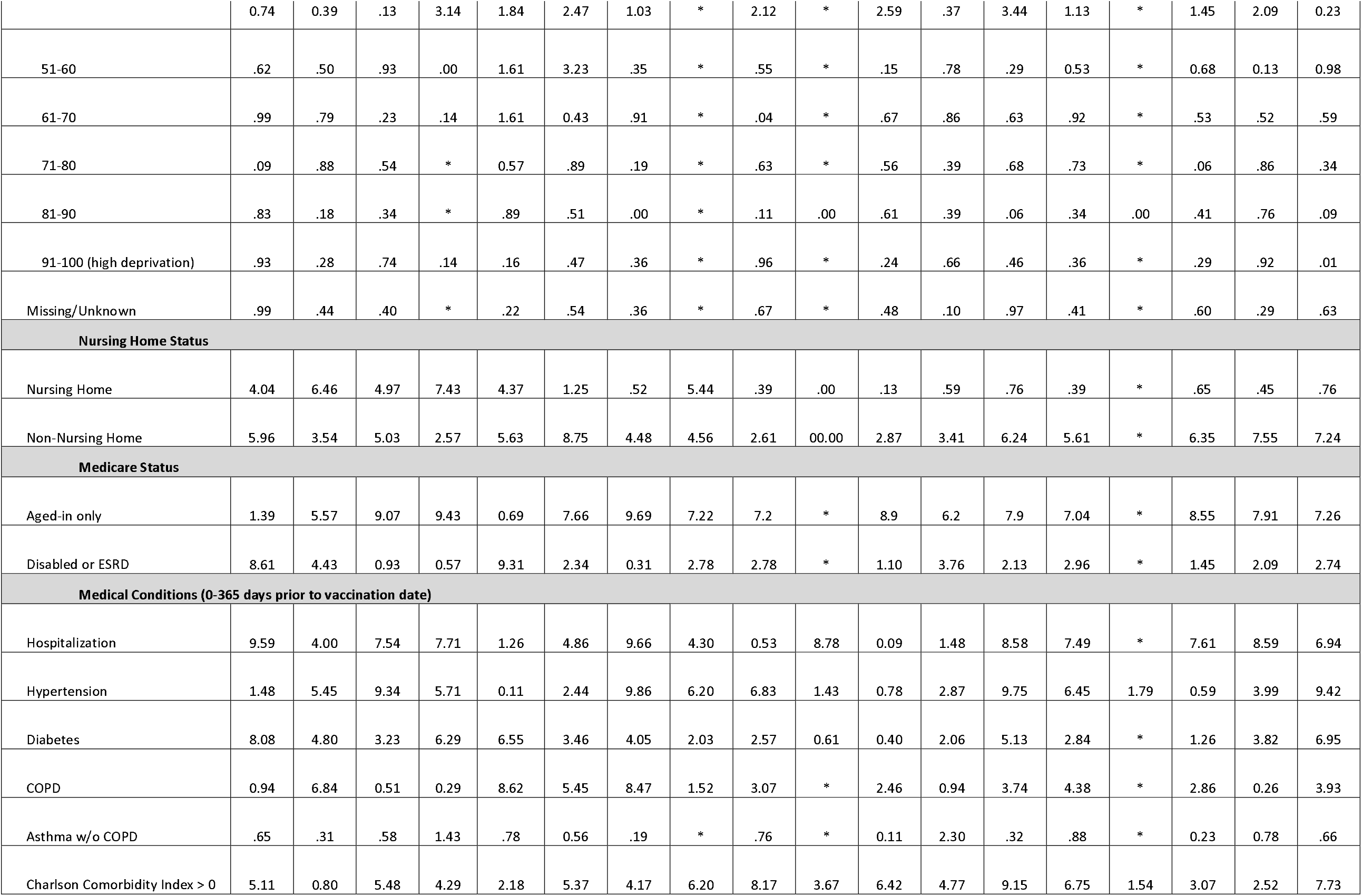
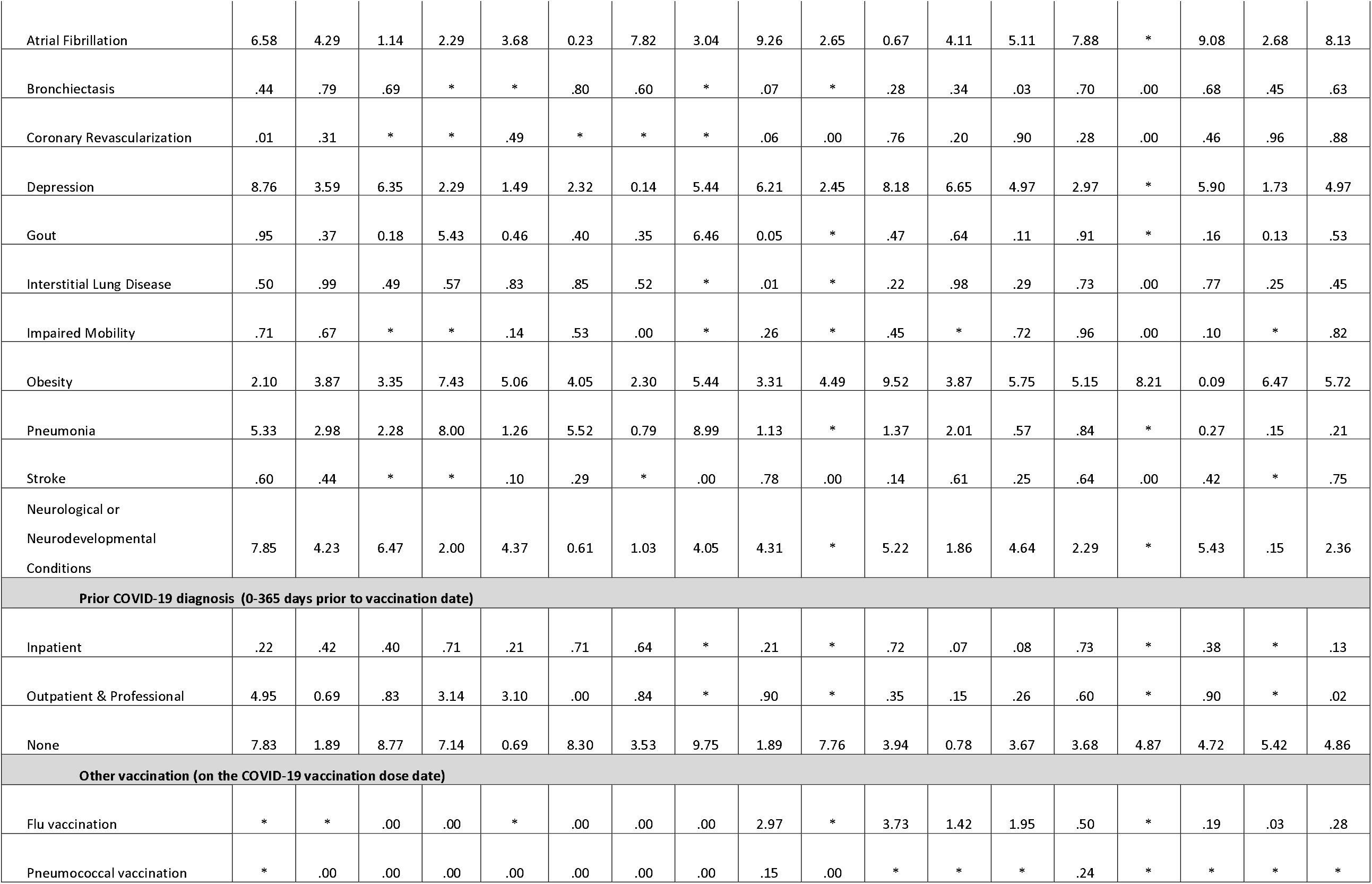
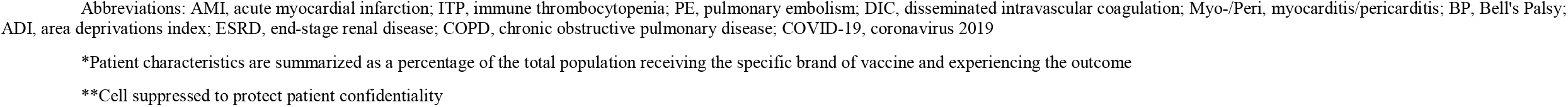
Descriptive summary of case characteristics for primary SCCS analysis for primary series and booster dose COVID-19 vaccinees, by vaccine brand and adverse event.

**Table 3.** Summary of MRR Case Adjudication Results and PPVs Associated with Adverse Events.

| Outcome and Final Case Classifications | Risk and Control Cases* | Risk Cases<sup>†</sup> | Control Cases<sup>††</sup> |
| --- | --- | --- | --- |
| **AMI** | **92** | **50** | **42** |
| Confirmed case | 35 | 20 | 15 |
| Probable | 37 | 21 | 16 |
| Possible | 15 | ** | ** |
| Not a case | ** | ** | ** |
| Unable to be determined | ** | ** | 0 |
| PPV (Confirmed + Probable)<sup>¶</sup> | 80.00%<br>(70.59%,<br>86.96%) | 85.42%<br>(72.83%,<br>92.75%) | 73.81%<br>(58.93%,<br>84.70%) |
| **PE** | **101** | **59** | **42** |
| Confirmed case | ** | ** | ** |
| Probable | ** | ** | ** |
| Possible | ** | ** | ** |
| Not a case | 46 | 29 | 17 |
| Unable to be determined | ** | ** | ** |
| PPV (Confirmed + Probable)<sup>¶</sup> | 45.74%<br>(36.04%,<br>55.78%) | 41.82%<br>(29.74%,<br>54.97%) | 51.28%<br>(36.20%,<br>66.13%) |
| **PE (IP)** | **42** | **23** | **19** |
| Confirmed case | ** | ** | ** |
| Probable | ** | ** | ** |
| Possible | ** | ** | ** |
| Not a case | ** | 0 | ** |
| Unable to be determined | 0 | 0 | 0 |
| PPV (Confirmed + Probable)<sup>¶</sup> | 83.33%<br>(69.40%,<br>91.68%) | 91.30%<br>(73.20%,<br>97.58%) | 73.68%<br>(51.21%,<br>88.19%) |
| **ITP** | **91** | **53** | **38** |
| Confirmed case | ** | ** | ** |
| Probable | ** | ** | ** |
| Possible | ** | ** | ** |
| Not a case | 66 | 39 | 27 |
| Unable to be determined | 16 | ** | ** |
| PPV (Confirmed + Probable)<sup>¶</sup> | 4.00%<br>(1.37%, 11.11%) | 4.65%<br>(1.28%, 15.46%) | 3.12%<br>(0.55%, 15.74%) |
| **DIC** | **90** | **48** | **42** |
| Confirmed case | 35 | 20 | 15 |
| Probable | 0 | 0 | 0 |
| Possible | 24 | 12 | 12 |
| Not a case | ** | ** | ** |
| Unable to be determined | ** | ** | ** |
| PPV (Confirmed) ¶ | 42.68%<br>(32.54%,<br>53.48%) | 46.51%<br>(32.51%,<br>61.08%) | 38.46%<br>(24.89%,<br>54.10%) |

Abbreviations: AMI, acute myocardial infarction; ITP, immune thrombocytopenia; PE, pulmonary embolism; DIC, disseminated intravascular coagulation; PPV, positive predictive value
\* Cases that occurred during either the risk or the control interval
\*\* Cell suppressed to protect patient confidentiality
† Cases that occurred during the risk interval
†† Cases that occurred during the control interval
¶ PPV Calculation excludes cases that we are unable to be determined/assigned a case classification based on MRR

For all six AEs included in either the primary series or booster studies, seasonality patterns varied, indicating the need for a seasonality adjustment comparing incidence rates during corresponding calendar months (data not shown). For the booster study, we observed the largest variation in seasonality patterns comparing the pre-pandemic (2018-2019) and pandemic (2020-2022) periods for PE, motivating the PE-specific post-hoc analysis adjusting for seasonality using event incidence rates during the pandemic study period (data not shown).

Descriptive analyses of cases in the primary series study found that AMI and DIC exhibited a high case fatality rate of 34% and 67%, respectively, indicating the need to adjust for curtailed observation time (eTable 3). AMI, inpatient PE, and DIC had a high proportion of cases with prior medically attended COVID-19 infection, ranging from 28% to 36% (eTable 4).

### 3.2 Medical Record Review

Table 3 summarizes results of medical record review (MRR) to verify outcomes by AE for the primary series study. MRR was conducted for a sample of cases with AMI, PE, ITP, and DIC events. AMI (PPV: 80.00% (95% CI: 70.59-86.96%)) and inpatient PE (PPV: 83.33% (95% CI: 69.40-91.68%)) claims-based definitions had the highest PPVs, indicating relatively accurate identification of cases. Comparatively, the all-care-setting PE (PPV: 45.74% (95% CI: 36.04-55.78%)) and DIC (PPV: 42.68% (95% CI: 32.54-53.48%)) event definitions had lower PPV estimates in identifying true disease cases. The all-care-setting ITP (PPV: 4.00% (95% CI:1.37-11.11%)) event definition used in the primary series study had the lowest PPV of all AEs, pointing to high misclassification of this event.

### 3.3 Inferential Results

Results from the primary analyses are described below for the primary series and booster studies. Secondary and exploratory analyses for the primary series study (eTable 7 and eTable 8, respectively) as well as secondary analyses for the booster study are included in the supplemental material (eTable 11).

#### Acute Myocardial Infarction (AMI)

AMI was evaluated in both primary series and booster studies. We did not find consistent results for AMI risk following administration of the BNT162b2 or mRNA-1273 vaccines in both primary series and booster studies (Figure 1).

**Figure 1.**
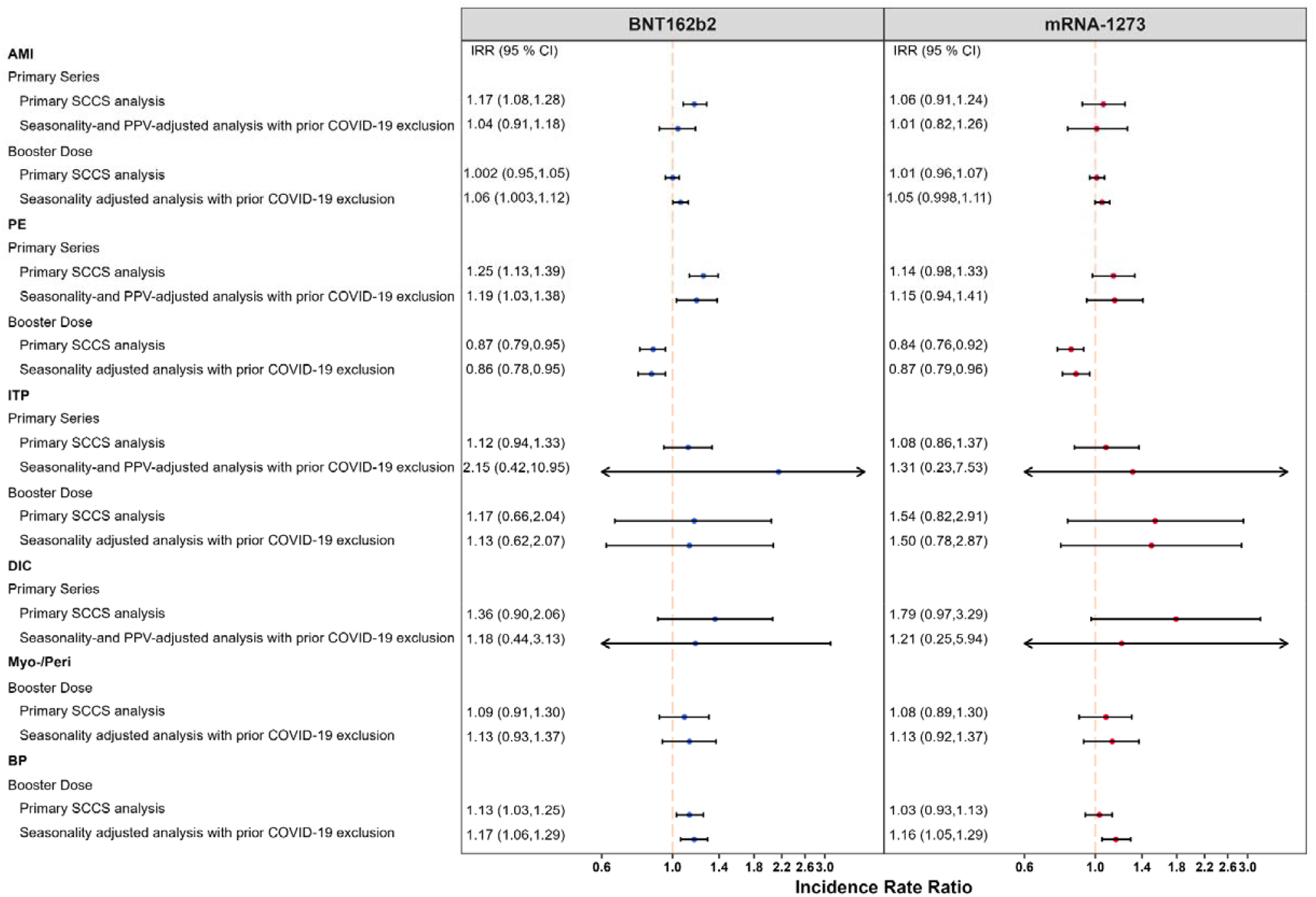
Summary of Self-Controlled Case Series (SCCS) Analysis with Post-Vaccine Control Interval Assessing the Incidence Rate Ratio (IRR) Comparing Risk and Control Periods of Acute Myocardial Infarction (AMI), Pulmonary Embolism (PE), Immune Thrombocytopenia (ITP), Disseminated Intravascular Coagulation (DIC), Myocarditis/Pericarditis (Myo-/Peri), and Bell ‘s Palsy (BP) Following a Primary Series or Booster COVID-19 Vaccine, Adjusting for Seasonality, Curtailed Observation Time, Interval-Specific PPV, and Excluding Cases with Evidence of Prior COVID-19. *Figure 1 displays incident rate ratios and corresponding 95% confidence intervals from eTables 5, 6, 9, and 10

We detected a small but statistically significant elevated risk of AMI following the BNT162b2 primary series (IRR=1.17, 95% CI: 1.08 to 1.28) (Figure 1). However, this effect was no longer statistically significant when accounting for seasonality, adjusting for outcome misclassification using MRR-derived PPVs, and excluding individuals with prior COVID-19 infection (IRR=1.04, 95% CI: 0.91 to 1.18). We did not observe evidence of elevated risk following the BNT162b2 booster dose (IRR=1.00, 95% CI: 0.95 to 1.05) in the primary analysis. After adjusting for seasonality and exclusion of cases with prior COVID-19 infection, a small but statistically significant elevation in risk was observed following the BNT162b2 booster dose (IRR=1.06, 95% CI: 1.003 to 1.12). In the primary analysis, AR per 100,000 doses following BNT162b2 primary series vaccination (AR: 22.91, 95% CI: 10.79 to 35.02, eTable 5) was substantially higher than post booster dose (AR: 0.15, 95% CI: -3.97 to 4.26, eTable 9).

There was no statistically significant increased risk of AMI following the mRNA-1273 primary series in both the primary analysis (IRR=1.06, 95% CI: 0.91 to 1.24) and the adjusted analysis accounting for seasonality, PPV adjustment, and excluding cases with prior COVID-19 infection (IRR=1.01, 95% CI: 0.82 to 1.26) (Figure 1). The estimates from the booster study were consistent with no statistically significant increase in risk observed in the primary analysis (IRR=1.01, 95% CI: 0.96 to 1.07) or the adjusted analysis (IRR=1.05, 95% CI: 0.998 to 1.11). Figure 2 presents a more detailed summary of all the AMI analyses and results in both studies.

**Figure 2.**
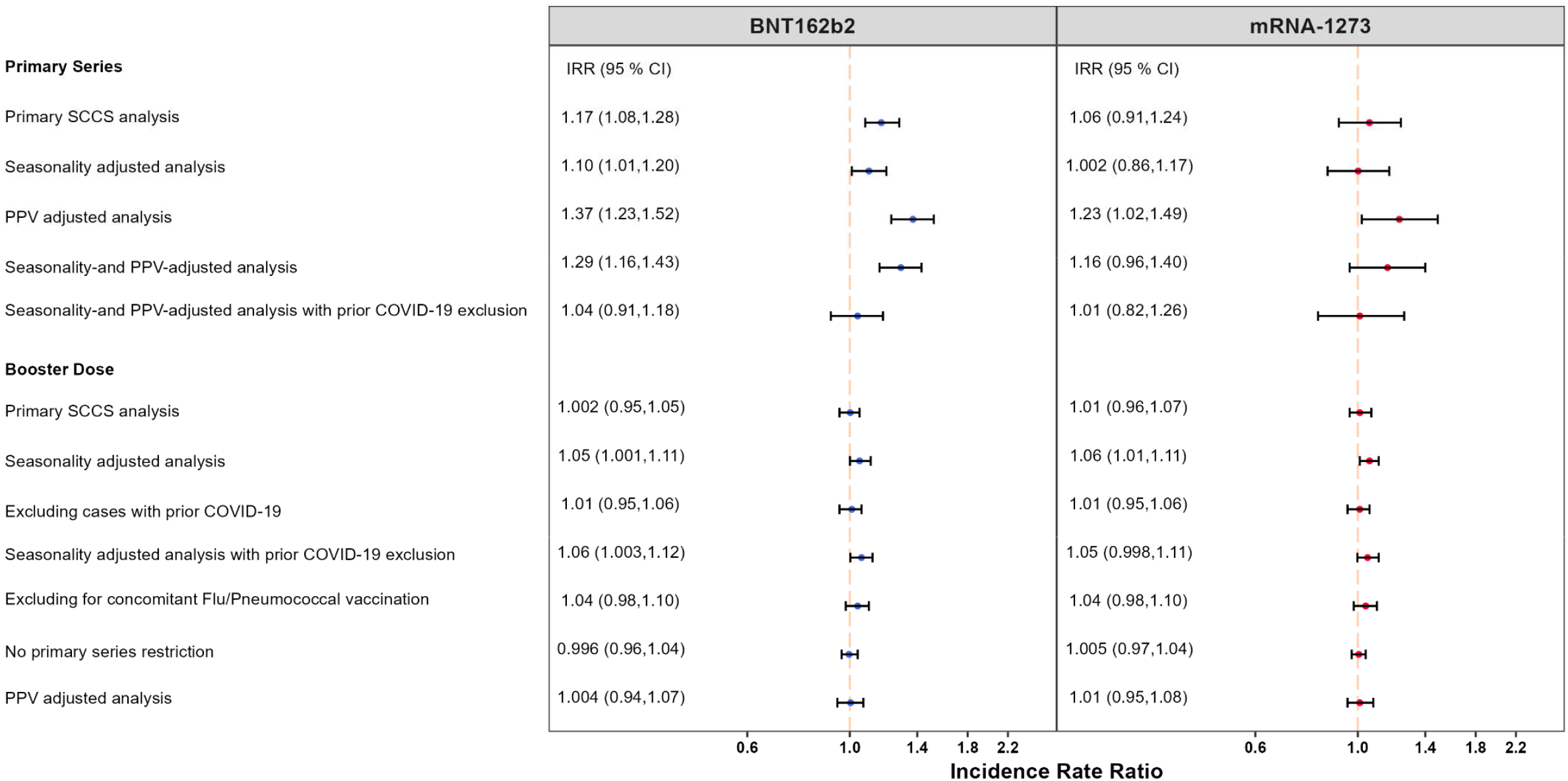
Summary of Self-Controlled Case Series (SCCS) Analysis with Post-Vaccine Control Interval Assessing the Incidence Rate Ratio (IRR) Comparing Risk and Control Periods of Acute Myocardial Infarction (AMI) Following a Primary Series or Booster COVID-19 Vaccine, Adjusting for Seasonality, Curtailed Observation Time, Interval-Specific PPV, and Excluding Cases with Evidence of Prior COVID-19.

#### Pulmonary Embolism (PE)

Inpatient PE was evaluated in both primary series and booster studies. We detected a small but statistically significant elevated risk of inpatient PE following BNT162b2 vaccination (IRR: 1.25, 95% CI: 1.13 to 1.39) in the primary series study, which remained significant in analyses adjusting for seasonality and PPV, and excluding individuals with prior COVID-19 infection (Figure 1). However, the booster study showed a statistically significant reduction in inpatient PE risk associated with BNT162b2 (IRR: 0.87, 95% CI: 0.79 to 0.95), which remained consistent after adjusting for seasonality and PPV, and excluding individuals with prior COVID-19 infection (IRR: 0.86, 95% CI: 0.78 to 0.95) and in most additional analyses (Figure 3). In the primary analysis, AR per 100,000 doses after primary series (AR: 7.04, 95% CI: 3.84 to 10.25, eTable 5) was higher than that following the booster dose (AR: -3.71, 95% CI: -5.99 to -1.44, eTable 9).

**Figure 3.**
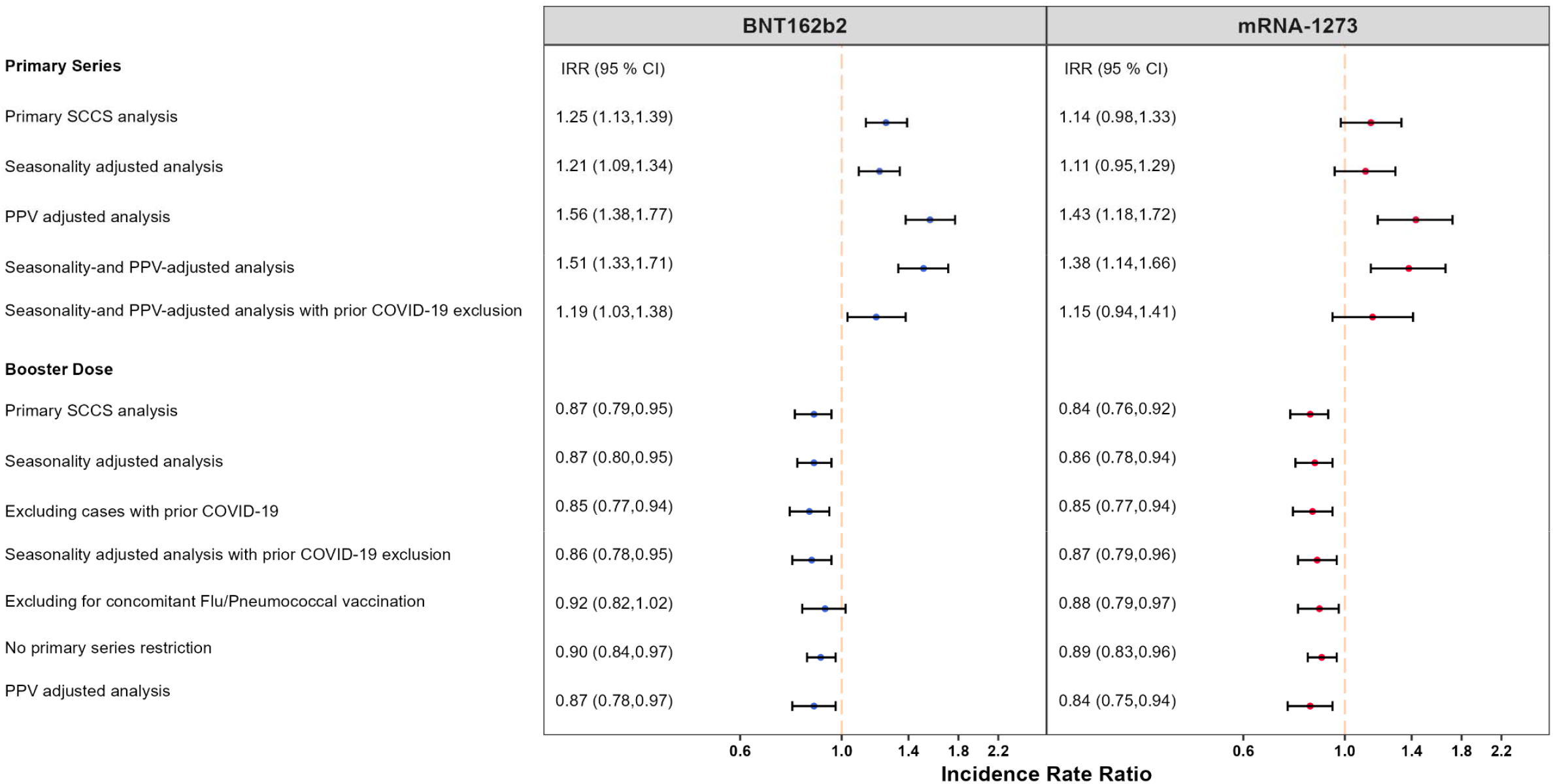
Summary of Self-Controlled Case Series (SCCS) Analysis with Post-Vaccine Control Interval Assessing the Incidence Rate Ratio (IRR) Comparing Risk and Control Periods of Pulmonary Embolism (PE) Following a Primary Series or Booster COVID-19 Vaccine, Adjusting for Seasonality, Curtailed Observation Time, Interval-Specific PPV, and Excluding Cases with Evidence of Prior COVID-19.

In the primary series study, we did not detect a statistically significant elevated risk of inpatient PE following mRNA-1273 in the primary analysis (IRR: 1.14, 95% CI: 0.98 to 1.33) or in the analysis adjusting for seasonality and PPV, and prior COVID-19 infection (IRR: 1.15, 95% CI: 0.94 to 1.41) (Figure 1). The booster study showed a statistically significant decrease in inpatient PE risk in the primary analysis (RR: 0.84, 95% CI: 0.76 to 0.92) and analyses adjusting for seasonality and PPV, and excluding individuals with prior COVID-19 infection (IRR: 0.87, 95% CI: 0.79 to 0.96) and in all other adjustments (Figure 3). In the primary analysis, AR per 100,000 doses after primary series (AR: 3.42, 95% CI: -0.46 to 7.30, eTable 5) was higher than that following the booster dose (AR: -4.08, 95% CI: -6.15 to -2.01, eTable 9).

Figure 5 presents a more detailed summary of all the BP analyses and results in both studies.

#### Bell ‘s Palsy (BP)

BP was evaluated in the booster study only. A small but statistically significant elevation in BP risk was detected following a booster dose of BNT162b2 vaccination (IRR: 1.13, 95% CI: 1.03 to 1.25), and it remained consistent across additional analyses such as adjustment for seasonality and prior COVID-19 infection exclusion (IRR: 1.17, 95% CI: 1.06 to 1.29) (Figure 5). There was no statistically significant elevation in BP risk after a booster dose of mRNA-1273 vaccination (RR: 1.03, 95% CI: 0.93 to 1.13), and it remained consistent across some additional analyses although a statistically significant result was detected for the primary analysis adjusted for seasonality and exclusion for prior COVID-19 infection (IRR: 1.16, 95% CI: 1.05 to 1.29) (Figure 5). AR per 100,000 doses post-BNT162b2 was (AR: 3.21, 95% CI: 0.74 to 5.69) larger than that post-mRNA-1273 (AR: 0.63, 95% CI: -1.72 to 2.98) (eTable 9).

#### Other Adverse Events (ITP, DIC, and Myo/Peri)

ITP was evaluated in both primary series and booster studies. We did not find a statistically significant increase in ITP risk in any of the analyses in either primary series or booster study after exposure to mRNA vaccines. Following primary series of BNT162b2 and mRNA-1273, the seasonality and PPV-adjusted analyses with prior COVID-19 exclusion resulted in IRR: 2.15 (95% CI: 0.42 to 10.95), and IRR: 1.31 (95% CI: 0.23 to 7.53), respectively. Following booster dose of BNT162b2 and mRNA-1273, the seasonality analyses with prior COVID-19 exclusion resulted in IRR: 1.13 (95% CI: 0.62 to 2.07) and IRR: 1.50 (95% CI: 0.78 to 2.87), respectively (Figure 7).

DIC was evaluated only in the primary series study, and we did not detect a statistically significant increase in DIC risk in the analyses for either vaccine. Following primary series of BNT162b2 and mRNA-1273, the seasonality and PPV-adjusted analyses with prior COVID-19 exclusion resulted in IRR: 1.18 (95% CI: 0.44 to 3.13) and IRR: 1.21 (95% CI: 0.25 to 5.94), respectively (Figure 6).

Myo/Peri was evaluated only in the booster study, and similarly we did not observe a statistically significant increase in Myo/Peri risk in the analyses for either vaccine. Following booster dose of BNT162b2 and mRNA-1273, the seasonality analyses with prior COVID-19 exclusion resulted in IRR: 1.13 (95% CI: 0.93 to 1.37) and IRR: 1.13 (95% CI: 0.92 to 1.37), respectively (Figure 4).

**Figure 4.**
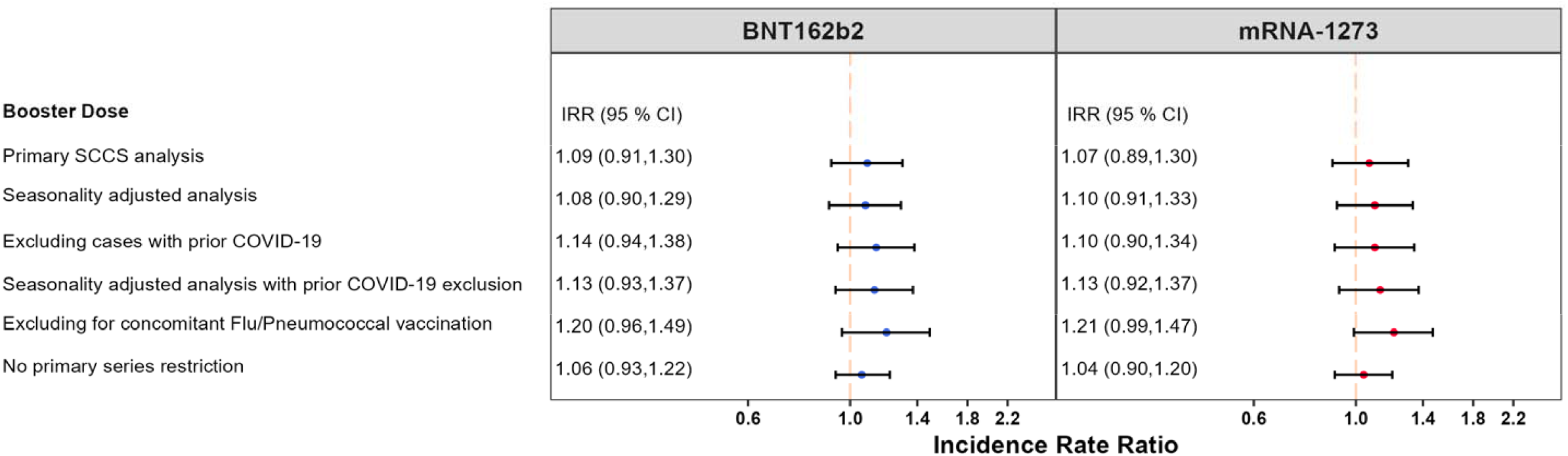
Summary of Self-Controlled Case Series (SCCS) Analysis with Post-Vaccine Control Interval Assessing the Incidence Rate Ratio (IRR) Comparing Risk and Control Periods of Myocarditis/Pericarditis (Myo-/Peri) Following a Booster COVID-19 Vaccine, Adjusting for Seasonality, Curtailed Observation Time, Interval-Specific PPV, and Excluding Cases with Evidence of Prior COVID-19.

**Figure 5.**
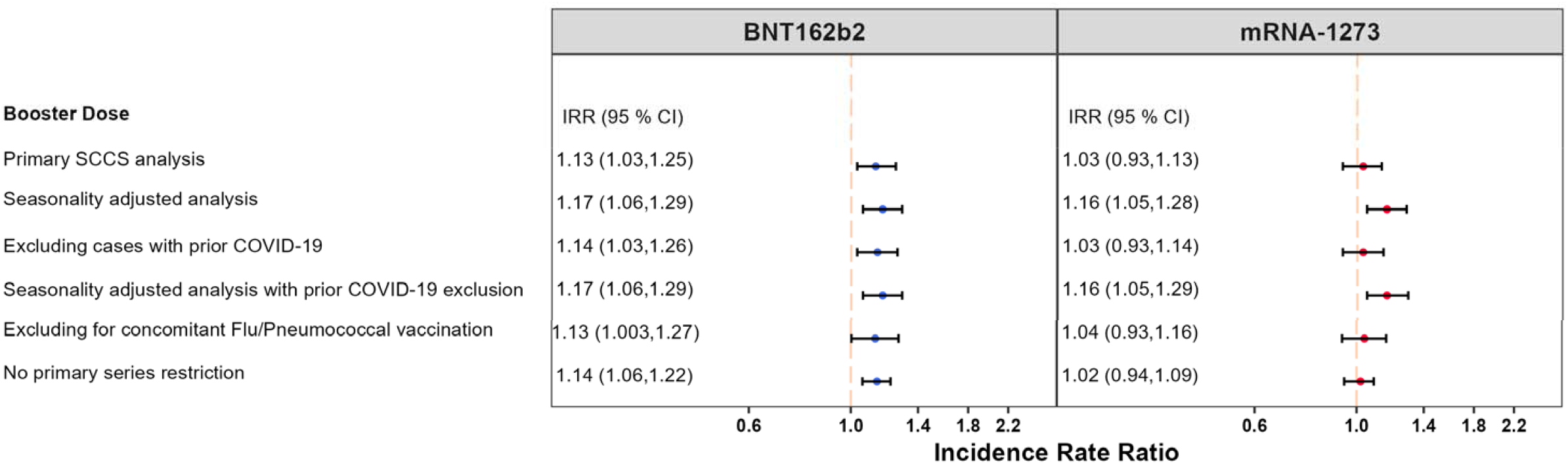
Summary of Self-Controlled Case Series (SCCS) Analysis with Post-Vaccine Control Interval Assessing the Incidence Rate Ratio (IRR) Comparing Risk and Control Periods of Bell ‘s Palsy (BP) Following a Booster COVID-19 Vaccine, Adjusting for Seasonality, Curtailed Observation Time, Interval-Specific PPV, and Excluding Cases with Evidence of Prior COVID-19.

**Figure 6.**
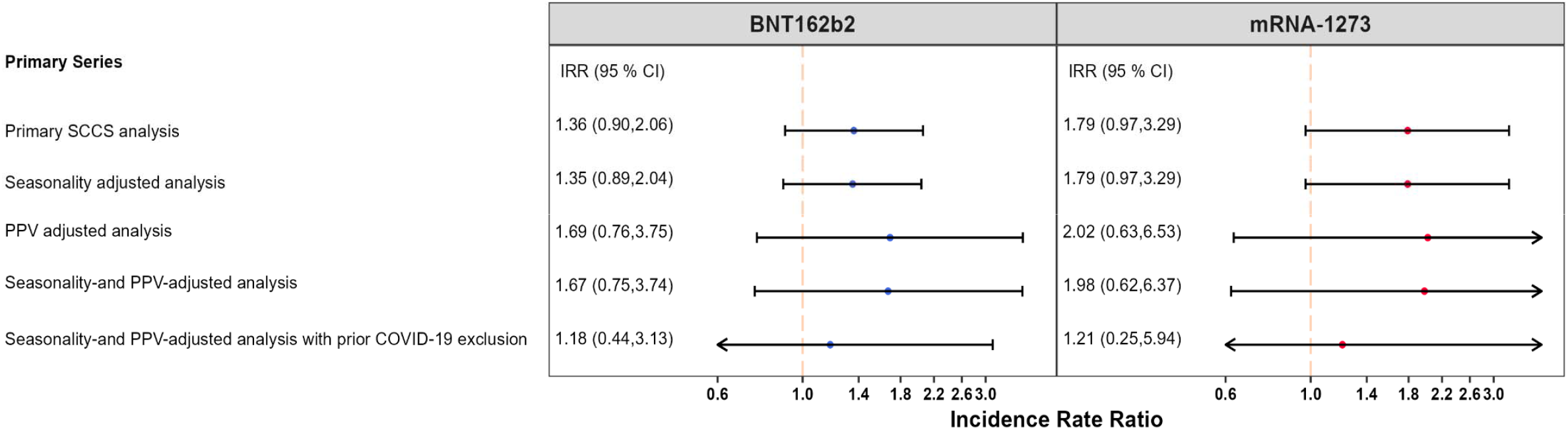
Summary of Self-Controlled Case Series (SCCS) Analysis with Post-Vaccine Control Interval Assessing the Incidence Rate Ratio (IRR) Comparing Risk and Control Periods of Disseminated Intravascular Coagulation (DIC) Following a Primary Series COVID-19 Vaccine, Adjusting for Seasonality, Curtailed Observation Time, Interval-Specific PPV, and Excluding Cases with Evidence of Prior COVID-19.

**Figure 7.**
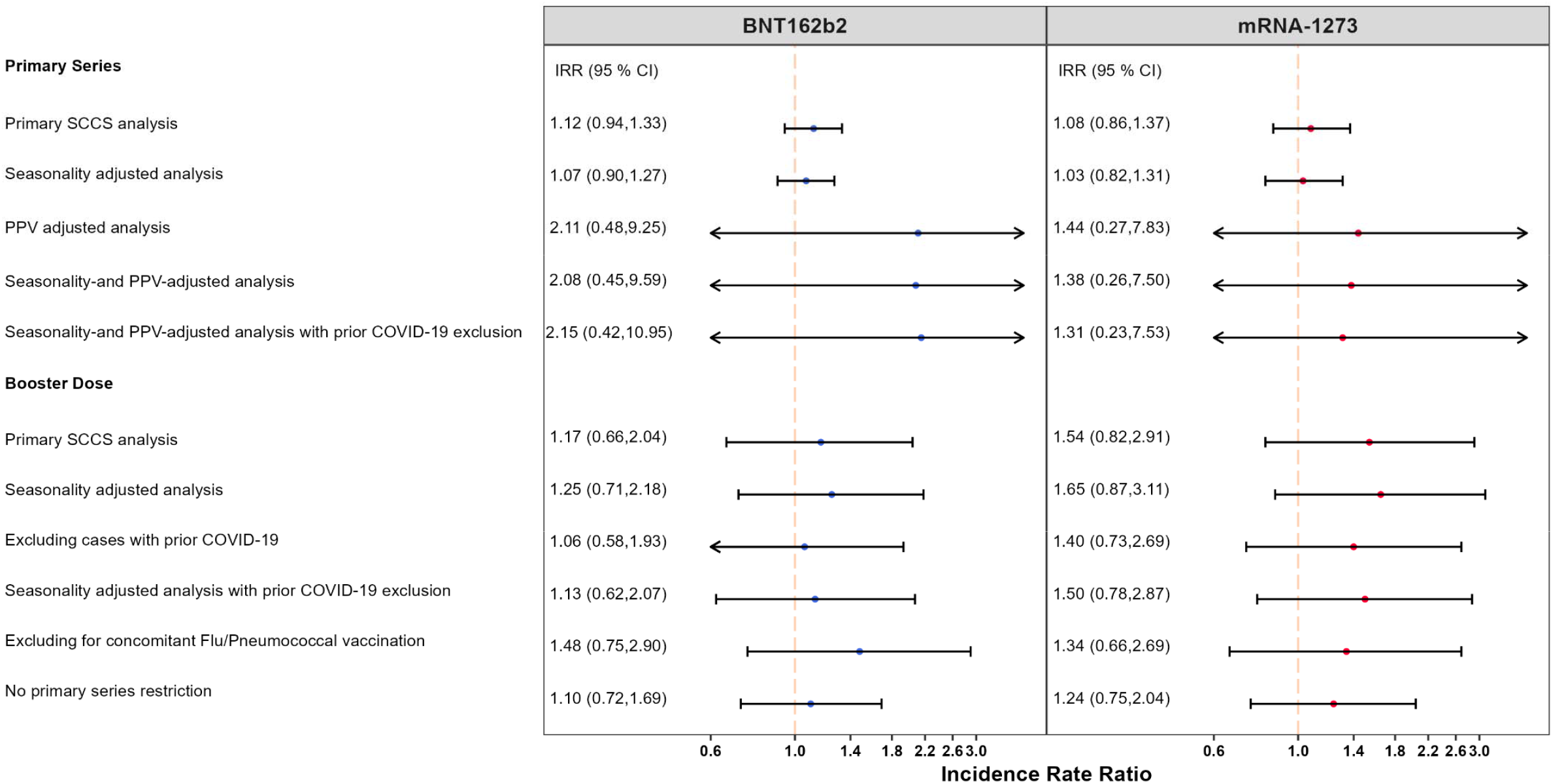
Summary of Self-Controlled Case Series (SCCS) Analysis with Post-Vaccine Control Interval Assessing the Incidence Rate Ratio (IRR) Comparing Risk and Control Periods of Immune Thrombocytopenia (ITP) Following a Primary Series or Booster COVID-19 Vaccine, Adjusting for Seasonality, Curtailed Observation Time, Interval-Specific PPV, and Excluding Cases with Evidence of Prior COVID-19.

Figures 4, 6, and 7 present a more detailed summary of all the analyses and results for ITP, DIC, and Myo/Peri, respectively, in both studies.

## 4 DISCUSSION

Of six AEs evaluated in two independent population-based studies including the U.S. elderly population, no statistically significant increase in risk was identified for ITP, DIC and Myo/Peri following COVID-19 mRNA vaccination for primary series and monovalent booster. These findings were robust to multiple analytic methods and adjustments.

The risk of AMI following COVID-19 mRNA vaccine primary series and booster doses was consistent across brands. The potential increased risk of AMI following the BNT162b2 primary series vaccine was reduced to a null effect after analytic adjustments. Following the booster BNT162b2 vaccine, the null effect from the primary analysis became a slightly elevated risk after analytic adjustments. The increased risk of AMI after BNT162b2 booster dose was small. In both the primary and adjusted analyses, there was no evidence of a statistically significantly increased AMI risk after exposure to the primary series and booster doses of mRNA-1273 vaccine.

There was not consistent evidence of an increased risk of inpatient PE following COVID-19 mRNA vaccination between the primary series and booster studies. Following the primary series of BNT162b2 vaccination we observed a small but statistically significant increase in inpatient PE risk that was robust to analytical adjustments; however, the risk following mRNA-1273 primary series was not statistically significantly elevated. In contrast, exposure to both mRNA vaccines booster doses showed a statistically significant protective effect against inpatient PE risk that was largely robust to analytical adjustments. The increased risk of PE following BNT162b2 primary series and its decreased risk following booster doses of both mRNA vaccines were both small. The protective effect following a booster dose when there was a small but elevated PE risk following the primary series would be unexpected if there were a true increased PE risk following COVID-19 mRNA vaccination. This adds to the level of uncertainty about an increased PE risk.

The primary series study did not evaluate BP risk since it did not signal in the signal detection study. The booster study showed a small but statistically significantly increased risk of BP after exposure to BNT162b2 vaccine which remained significant after analytic adjustments. The small increased risk of BP following mRNA-1273 booster dose was only statistically significant in the adjusted analysis.

One potential explanation for some of the statistically significant results associated with AMI, PE, and BP after exposure to the COVID-19 mRNA vaccines in these two studies could be attributed to the fact that the studies implemented multiple analyses and designs and they have a large sample size which could increase the probability of detecting statistically significant but not necessarily clinically significant results.

The results from both primary series and booster studies contribute to the safety profile of COVID-19 mRNA vaccines.^19,24-30^ and they are largely consistent with the results of other studies. Using an SCCS study design, Jabagi, Botton, and Bertrand (2022) did not observe an increased risk of AMI, PE, and stroke within 14 days after BNT162b2 vaccine doses among French vaccine recipients aged 75 years or older.^26^ Whiteley et al. (2022) found lower risk of arterial and venous thrombotic events after BNT162b2 COVID-19 vaccination in those aged 70 years and older.^30^ Two UK and Scotland studies using the SCCS study design also found no evidence for elevated risk of thrombocytopenia, venous thromboembolism, myocardial infarction, or hemorrhagic events following BNT162b2 vaccine administration in the general population.^19,28^ A Danish study on frontline workers found no association of thrombosis and thrombotic events with the BNT162b2 vaccine.^25^ Welsh et al. (2021) assessed thrombocytopenia cases (including ITP) reported to the Vaccine Adverse Event Reporting System (VAERS) and found that the risk of ITP following mRNA vaccines did not exceed expected historical rates.^29^ Berild et al. (2022) found increased rates of several thromboembolic and thrombocytopenia outcomes following mRNA vaccines, however this result was not robust to sensitivity analyses and was smaller compared to post vaccination risk following ChAdOx1 vaccines.^31^ Similarly, a number of case reports and studies have indicated a small but elevated risk of Myo/Peri in young males 16-24 years following COVID-19 mRNA vaccination; however, no strong evidence has indicated an elevated risk among individuals aged 65 years and older.^32,33^

The evidence of PE risk following COVID-19 mRNA vaccination in our studies is mixed, and this observation is similarly reflected in the current literature. Despite case reports documenting the occurrence of PE cases following COVID-19 vaccination with a proposed etiology of inflammatory response in susceptible patients,^34,35^ multiple studies suggest a lack of evidence for a strong association between elevated risk of venous thromboembolism events as well as PE, specifically following the BNT162b2 vaccine.^19,25,26,28,30,36^ This elevated risk has been more commonly associated with the ChAdOx1 nCoV-19 vaccine.^30^ While Burn et al. (2022) detected a slightly elevated PE risk following both vaccines, the risk was substantially higher following COVID-19 infection. ^36^ Several studies have similarly shown an increased PE risk following COVID-19 infection.^20,24^ Our investigation into the seasonality of PE during the pandemic suggests a strong correlation between spikes in COVID-19 infection and PE occurrence. When excluding PE cases with evidence of prior medically attended COVID-19 infection, the increased PE risk following mRNA vaccination attenuated but remained elevated. This however conflicts with the protective PE effect observed following booster doses for both mRNA vaccines which suggests that the exclusion of cases with evidence of prior COVID-19 infection might not have completely accounted for this effect. Taken together, these observational studies do not provide conclusive evidence for increased PE risk following COVID-19 mRNA vaccination.

The elevated BP risk observed following both COVID-19 mRNA vaccines booster dose is not entirely consistent with the literature. Wan et al. (2022) did not find a statistically significant increase in BP risk associated with BNT162b2 vaccination in a study conducted in China.^37^ While several case reports have also cited BP cases following COVID-19 mRNA vaccines, Renoud et al. (2021) suggest that the reporting rate of BP after COVID-19 mRNA vaccination is comparable to other viral vaccines from a disproportionality analysis conducted in the World Health Organization (WHO) pharmacovigilance database.^38^ Shemer (2021) did not identify a statistically significant association between BP risk and COVID-19 mRNA vaccination.^39^ Tamaki (2021) suggests that the risk of BP is seven times more likely after SARS-Cov-2 infection than after COVID-19 vaccination.^40^ The risk of AMI, PE and BP secondary to SARS-CoV-2 infections were reported in several studies to be substantially higher than post-vaccination risk estimates.^40,41^ Challenges in confirming prior COVID-19 infection in vaccinees, especially near the time of vaccination, may complicate our ability to obtain accurate estimate of some of these outcomes.

These two observational studies have several strengths. The SCCS study design inherently adjusts for potential time-invariant confounders which may draw from between-individual comparisons. The large size of the CMS Medicare population provides more power for the study to evaluate rare AEs with more precision. The Medicare database is a large, population-based database containing information on beneficiaries ‘ demographics and longitudinal information on health care services utilization across care settings, thus more comprehensively capturing people ‘s baseline health conditions and across time. Further, since a large proportion of the U.S. elderly population is enrolled in Medicare and beneficiary attrition is minimal once eligible, our findings are highly generalizable to the U.S. population aged 65 years and older. While implementation of multiple study designs and analytic methods in the observational studies examines the robustness of the risk estimates, it can also increase the likelihood of detecting statistically significant results due to chance alone.

Potential vaccine exposure misclassification cannot be ruled out in the studies given the current evidence with respect to under-reporting of COVID-19 vaccine administration in medical claims data sources.^42^ However, the impact of exposure misclassification on the risk estimates may not be large since the study population in both studies was comprised of only vaccinated individuals. Outcome misclassification cannot be ruled out either especially in regards to the use of ‘rule-out diagnosis ‘ in administrative claims. Also, MRR results were not available at the time of the study to confirm the outcome status for all AEs in these studies; and some outcomes for which MRR was conducted had low PPV. Misspecification of risk and control intervals could also bias the estimates in either direction. Finally, since residual and unmeasured confounding in observational studies cannot be fully ruled out, the results carry a certain level of uncertainty.

In these two studies of the U.S. elderly we did not find an increased risk for AMI, ITP, DIC, and Myo/Peri; the results were not consistent for PE; and there was a small elevated risk of BP after exposure to COVID-19 mRNA vaccines. These results support the safety profile of COVID-19 mRNA vaccines administered to the U.S. elderly and are consistent with the conclusion that the benefits of COVID-19 vaccination outweigh the risks of disease.

## Data Availability

All data described in the present study is not available to protect patient confidentiality.

## ACKNOWLEDGEMENTS

Funding was provided by the US Food and Drug Administration. We would like to thank Purva Shah, Amei Hao, Yeerae Kim, Katie Matuska, and Mahasweta Mitra for providing statistical programming and writing support.

## Tables and Figures for

Evaluation of Potential Adverse Events Following COVID-19 mRNA Vaccination Among Adults Aged 65 Years and Older: A Self-Controlled Study in the U.S.Figure 2 presents a more detailed summary of all the AMI analyses and results in both studies.

