## Supplemental Materials for "Evaluation of Potential Adverse Events Following COVID-19 mRNA Vaccination Among Adults Aged 65 Years and Older: A Self-Controlled Study in the U.S"

Note: All secondary and exploratory analyses are not reported in the supplemental tables for brevity.

**eTable 1. Summary of analytical plan and study specifications**

| **Analysis Specifications** | **Summary** |
| --- | --- |
| **Overview** | The analysis aims to assess the association of Pfizer-BioNTech (BNT162b2), and Moderna (mRNA-1273) vaccine administration with the six adverse events that met the threshold of a statistical signal in the RCA in the Medicare population, accounting for potential sources of confounding using self-controlled study designs. |
| **Data Sources** | We used Medicare claims and enrollment files from the Centers for Medicare and Medicaid Services (CMS) Medicare Shared Systems Data (SSD). |
|  | Demographics and information on death is derived from the enrollment databases. Information on vaccinations, health covariates, preventive services, and outcomes are derived from Medicare Part A (inpatient) and Part B (outpatient and community settings) and Part D (prescription) claims. |
|  | Beneficiaries' nursing home residence information was assessed using assessment data from the Minimum Data Set (MDS). |
| **Study Period** | Primary Series COVID-19 Vaccinations: December 11, 2020 - April 16, 2021 (AMI), April 30, 2021 (PE, DIC), and May 7, 2021 (ITP) |
|  | Booster Dose COVID-19 Vaccinations: August 12, 2021 – April 30, 2022 (AMI, PE, ITP), May 7, 2022 (Myo/Peri), May 14, 2022 (BP) |
| **Exposures** | BNT162b2, and mRNA-1273 COVID-19 vaccine receipt was identified using Current Procedural Terminology (CPT®)/Healthcare Common Procedure Coding System (HCPCS) codes or National Drug Codes (NDCs) in any care setting. |
| **Observation Period** | For the primary self-controlled case series (SCCS) analysis, the observation period extends from the vaccination date through 90 days after vaccination. Beneficiaries are censored when they disenroll, receive a subsequent booster dose (only for the booster dose study), die or at the end of the study. For the secondary SCCS analysis and the exploratory self-controlled risk interval (SCRI) analysis, the observation period starts from the first day of the pre-vaccination control interval and extends to day 90 after vaccination. The pre-vaccination control interval is defined as the interval with the same length as the risk interval ending 15 days prior to the date of the primary series or the booster dose COVID-19 vaccination. |
| **Risk Interval and Control Interval** | For the SCCS analyses, risk interval is defined as the time during which excess risk is hypothesized following each COVID-19 vaccine dose for the adverse events based on biological plausibility and clinical input. The control interval is made of all remaining time in the observation period. |
| **Population** | Eligible study population included those who received at least one dose of the primary series COVID-19 vaccine or the booster dose, experienced an incident adverse event in the observation period, and met the following inclusion/exclusion criteria: |
|  | (i) Enrolled in Parts A (hospital insurance) and Part B (medical insurance) fee-for-service (FFS) from the clean window prior to the occurrence of the adverse event through the study observation period or death |
|  | (ii) Received at least one dose of the primary series COVID-19 vaccine or a booster dose during the vaccination period of interest |
|  | (iii) Had a record of the adverse event diagnosis during the study observation period |
|  | (iii) At least 65 years of age at the time of COVID-19 vaccination |
|  | (iv) Contributed follow-up time to both risk and control intervals (except beneficiaries who died and could not contribute time to the control interval) |
|  | (v) Did not have a diagnosis of the adverse event of special interest (AESI) during the AESI-specific clean window |
|  | ***Exclusions implemented for the primary series population only:*** |
|  | (v) Had <17 days between first and second doses |
|  | (vi) Received different COVID-19 vaccines for the first and second dose observation |
|  | ***Exclusions implemented for the booster dose population only:*** |
|  | (v) Individuals vaccinated with multiple brands on the same day or multiple vaccination within 3 days of each other |
|  | (vi) Received different COVID-19 vaccines for the booster dose compared to the primary series vaccine dose. Additional, adjusted analysis was conducted which did not require beneficiaries to have a primary series vaccination. |
| **Outcomes** | Incident occurrences of acute myocardial infarction (AMI), pulmonary embolism (PE), disseminated intravascular coagulation (DIC), immune thrombocytopenia (ITP), Bell’s Palsy (BP), and myocarditis/pericarditis (Myo-/Peri)were identified with International Classification of Diseases, Tenth Revision, Clinical Modification (ICD-10-CM) codes. The first occurrence of the event was defined as an incident case if no event was recorded during the preceding pre-defined interval, i.e., the clean window (eTable 2) |
| **Covariates** | The following covariates were assessed descriptively: |
|  | (i) Demographics were assessed at the time of COVID-19 vaccination receipt, including age, sex, race/ethnicity, region, urban/rural, dual eligibility status, area deprivation index (ADI), nursing home residency status |
|  | (ii) Influenza vaccine receipt was assessed at the same time as the COVID-19 vaccine. Other vaccines (i.e., influenza and pneumococcal) were assessed during the observation period |
|  | (iv) History of a medically attended COVID-19 infection, defined by the ICD-10-CM codes U07.1 (COVID-19) were assessed using all available data |
|  | (vii) Risk factors for the adverse event were assessed in the 365 days prior to index date |
| **Additional Confounding Control** | The following adjustments were performed independently and concurrently to assess their impact on the primary analysis results: |
|  | We controlled for seasonal variation in adverse event occurrence in the primary SCCS, secondary SCCS, and exploratory SCRI analysis, using outcome incidence estimated from 2018 (AMI) and 2019 (ITP, DIC, PE, Myo-/Peri, BP). |
|  | With COVID-19 infection hypothesized as a potential risk factor for the adverse events assessed in the study, we excluded cases with evidence of prior medically attended COVID-19 infection from the primary SCCS analysis. |
|  | We minimized potential outcome misclassification confounding by performing quantitative bias assessments informed by medical record review (MRR) in the primary SCCS, secondary SCCS, and exploratory SCRI analysis. |
| **Medical Record Review (MRR) and Quantitative Bias Assessment (QBA)** | MRR: To validate our claims-based adverse event case definition, we retrieved a sample of up to 100 cases’ medical records for each outcome, which underwent an adjudication process by board-certified clinicians. Cases were identified as true cases, non-cases, and potentially indeterminate using pre-determined clinical definitions. |
|  | Quantitative bias assessment: A multiple imputation approach was used, where simulated datasets were created based on chart-confirmed cases by sampling with probability equal to the positive predictive value derived from the medical record review of the adjudicated cases. The conditional Poisson regression was repeated on the simulated datasets and risk estimates pooled across the simulated datasets. |
| **Statistical Analysis** | Rate ratios (RR), the ratio of the incidence in the exposure risk periods relative to the incidence in control periods, were estimated. |
|  | We also calculated absolute risk (AR), estimated as the number excess adverse events per 100,000 doses or person-years, if no vaccine was administered. |

**eTable 2. Adverse Events of Interest (AE), settings, clean windows, and risk windows**

| **Adverse Events** | **Care Setting** | **Clean Window** | **Risk Window** |
| --- | --- | --- | --- |
| Acute Myocardial Infarction (AMI) | IP | 365 days* | 1-28 days† |
| Pulmonary Embolism† (PE) | IP | 365 days* | 1-28 days† |
| Disseminated Intravascular Coagulation (DIC) | IP, OP-ED | 365 days* | 1-28 days† |
| Immune Thrombocytopenia (ITP) | IP, OP/PB (for the primary series study) | 365 days* | 1-42 days† |
|  | IP -Primary Diagnosis (for the booster dose study) |  |  |
| Bell’s Palsy (BP) | IP, OP/PB | 183 days* | 1-42 days† |
| Myocarditis/Pericarditis (Myo-/Peri) | IP, OP/PB | 365 days* | 1-21 days† |

* Selection of clean window duration is based on clinician input

† Selection of risk window based on literature review and informed by clinician input

IP = inpatient, OP-ED = outpatient emergency department, OP/PB = outpatient or provider

**eTable 3. Summary of primary SCCS analysis deaths during the observation period, by outcome, vaccine brand, and interval for the primary series COVID-19 study population**

| **Brand** | **Interval when outcome occurred** | **# of Cases** | **Deaths** | | **Deaths in Risk Interval** | | **Deaths in Risk Interval Before Accruing Control Time** | | **Deaths in the Control Interval** | |
| --- | --- | --- | --- | --- | --- | --- | --- | --- | --- | --- |
|  |  |  | **#** | **% of Cases** | **#** | **% of Cases** | **#** | **% of Cases** | **#** | **% of Cases** |
| **AMI** |  |  |  |  |  |  |  |  |  |  |
| Overall | | 3,653 | 1,249 | 34.19% | 606 | 16.59% | 582 | 15.93% | 643 | 17.60% |
| BNT162b2 | Control | 1,189 | 274 | 23.04% | ** | ** | - | - | ** | ** |
| BNT162b2 | Risk | 1,594 | 694 | 43.54% | 474 | 29.74% | 457 | 28.67% | 220 | 13.80% |
| mRNA-1273 | Control | 356 | 84 | 23.60% | ** | ** | - | - | ** | ** |
| mRNA-1273 | Risk | 514 | 197 | 38.33% | 131 | 25.49% | 125 | 24.32% | 66 | 12.84% |
| **PE (IP)** |  |  |  |  |  |  |  |  |  |  |
| Overall | | 2,470 | 566 | 22.91% | 226 | 9.15% | 200 | 8.10% | 340 | 13.77% |
| BNT162b2 | Control | 720 | 118 | 16.39% | ** | ** | - | - | ** | ** |
| BNT162b2 | Risk | 964 | 287 | 29.77% | 156 | 16.18% | 145 | 15.04% | 131 | 13.59% |
| mRNA-1273 | Control | 315 | 51 | 16.19% | ** | ** | - | - | ** | ** |
| mRNA-1273 | Risk | 471 | 110 | 23.35% | 66 | 14.01% | 55 | 11.68% | 44 | 9.34% |
| **ITP** |  |  |  |  |  |  |  |  |  |  |
| Overall | | 1,085 | 60 | 5.53% | 34 | 3.13% | 33 | 3.04% | 26 | 2.40% |
| BNT162b2 | Control | 196 | ** | ** | 0 | 0.00% | - | - | ** | ** |
| BNT162b2 | Risk | 472 | 39 | 8.26% | 25 | 5.30% | ** | ** | 14 | 2.97% |
| mRNA-1273 | Control | 99 | ** | ** | ** | ** | - | - | ** | ** |
| mRNA-1273 | Risk | 318 | 16 | 5.03% | ** | ** | ** | ** | ** | ** |
| **DIC** |  |  |  |  |  |  |  |  |  |  |
| Overall | | 254 | 170 | 66.93% | 92 | 36.22% | 83 | 32.68% | 78 | 30.71% |
| BNT162b2 | Control | 65 | 30 | 46.15% | ** | ** | - | - | ** | ** |
| BNT162b2 | Risk | 110 | 84 | 76.36% | ** | ** | 61 | 55.45% | ** | ** |
| mRNA-1273 | Control | 27 | 12 | 44.44% | 0 | 0.00% | - | - | 12 | 44.44% |
| mRNA-1273 | Risk | 52 | 44 | 84.62% | 29 | 55.77% | 22 | 42.31% | 15 | 28.85% |

*(-) Statistic is not applicable for the given group*

*** Cell suppressed to protect patient confidentiality*

**eTable 4. Summary of cases with prior COVID before the adverse event date for the primary series and booster dose COVID-19 study population**

| **Outcome** | **BNT162b2** | | | | | | **mRNA-1273** | | | | | |
| --- | --- | --- | --- | --- | --- | --- | --- | --- | --- | --- | --- | --- |
|  | **Risk Cases** | | | **Control Cases** | | | **Risk Cases** | | | **Control Cases** | | |
|  | **All** | **With Prior COVID** | | **All** | **With Prior COVID** | | **All** | **With Prior COVID** | | **All** | **With Prior COVID** | |
|  | **#** | **#** | **%** | **#** | **#** | **%** | **#** | **#** | **%** | **#** | **#** | **%** |
| **Primary Series Study** | | | | | | | | | | | | |
| AMI | 1594 | 671 | 42.10% | 1189 | 335 | 28.17% | 514 | 160 | 31.13% | 356 | 84 | 23.60% |
| PE (IP) | 964 | 364 | 37.76% | 720 | 172 | 23.89% | 471 | 125 | 26.54% | 315 | 43 | 13.65% |
| ITP | 472 | 63 | 13.35% | 196 | 33 | 16.84% | 318 | 32 | 10.06% | 99 | ** | ** |
| DIC | 110 | 42 | 38.18% | 65 | 22 | 33.85% | 52 | 22 | 42.31% | 27 | ** | ** |
| **Booster Dose Study** | | | | | | | | | | | | |
| AMI | 2,459 | 321 | 13.05% | 5,642 | 762 | 13.51% | 2,422 | 293 | 12.10% | 5,519 | 648 | 11.74% |
| PE | 724 | 123 | 16.99% | 1,898 | 294 | 15.49% | 657 | 88 | 13.39% | 1,806 | 267 | 14.78% |
| ITP | 24 | ** | ** | 25 | ** | ** | 22 | ** | ** | 17 | 0 | 0.00% |
| Myo-/Peri | 166 | 18 | 10.84% | 517 | 79 | 15.28% | 144 | 12 | 8.33% | 468 | 48 | 10.26% |
| BP | 827 | 56 | 6.77% | 847 | 60 | 7.08% | 752 | 48 | 6.38% | 842 | 55 | 6.53% |

*** Cell suppressed to protect patient confidentiality*

**eTable 5. Summary of incidence rate ratio and attributable risk of AMI, inpatient PE, ITP, and DIC in primary SCCS analysis for the primary series COVID-19 vaccinations with post-vaccine control interval, adjusting for event-dependent observation time**

|  | **BNT162b2** | | | | | **mRNA-1273** | | | | |
| --- | --- | --- | --- | --- | --- | --- | --- | --- | --- | --- |
| **AMI** |  |  |  |  |  |  |  |  |  |  |
| ***Conditional Poisson Regression*** |  |  |  |  |  |  |  |  |  |  |
| *Cases in Risk Window* | 1,594 | | | | | 514 | | | | |
| *Cases in Control Window* | 1,189 | | | | | 356 | | | | |
| *Person-Days in Risk Window* | 119,446 | | | | | 41,023 | | | | |
| *Person-Days in Control Window* | 87,888 | | | | | 25,161 | | | | |
| *Model Results* | IRR | 95% CI | | P-Value | SE | IRR | 95% CI | | P-Value | SE |
|  | 1.17 | 1.08 | 1.28 | <0.01 | 0.04 | 1.06 | 0.91 | 1.24 | 0.42 | 0.08 |
| ***Attributable Risk*** |  |  |  |  |  |  |  |  |  |  |
| *# of Eligible Vaccinated* | 535,252 | | | | | 205,350 | | | | |
| *# of Vaccines* | 1,020,071 | | | | | 390,303 | | | | |
| *Person-Years* | 68,689 | | | | | 29,540 | | | | |
| *Attributable Risk per 100,000 Doses* | AR | 95% CI | | P-Value | SE | AR | 95% CI | | P-Value | SE |
|  | 22.91 | 10.79 | 35.02 | <0.01 | 6.18 | 7.94 | -11.50 | 27.39 | 0.42 | 9.92 |
| *Attributable Risk per 100,000 Person-Years* | AR | 95% CI | | P-Value | SE | AR | 95% CI | | P-Value | SE |
|  | 340.20 | 160.31 | 520.10 | <0.01 | 91.79 | 104.96 | -151.97 | 361.88 | 0.42 | 131.09 |
| **PE (IP)** |  |  |  |  |  |  |  |  |  |  |
| ***Conditional Poisson Regression*** |  |  |  |  |  |  |  |  |  |  |
| *Cases in Risk Window* | 964 | | | | | 471 | | | | |
| *Cases in Control Window* | 720 | | | | | 315 | | | | |
| *Person-Days in Risk Window* | 74,248 | | | | | 38,846 | | | | |
| *Person-Days in Control Window* | 60,910 | | | | | 25,919 | | | | |
| *Model Results* | IRR | 95% CI | | P-Value | SE | IRR | 95% CI | | P-Value | SE |
|  | 1.25 | 1.13 | 1.39 | <0.01 | 0.05 | 1.14 | 0.98 | 1.33 | 0.08 | 0.08 |
| ***Attributable Risk*** |  |  |  |  |  |  |  |  |  |  |
| *# of Eligible Vaccinated* | 1,458,018 | | | | | 910,705 | | | | |
| *# of Vaccines* | 2,772,818 | | | | | 1,737,779 | | | | |
| *Person-Years* | 188,824 | | | | | 132,195 | | | | |
| *Attributable Risk per 100,000 Doses* | AR | 95% CI | | P-Value | SE | AR | 95% CI | | P-Value | SE |
|  | 7.04 | 3.84 | 10.25 | <0.01 | 1.64 | 3.42 | -0.46 | 7.30 | 0.08 | 1.98 |
| *Attributable Risk per 100,000 Person-Years* | AR | 95% CI | | P-Value | SE | AR | 95% CI | | P-Value | SE |
|  | 103.41 | 56.35 | 150.48 | <0.01 | 24.01 | 44.97 | -5.99 | 95.93 | 0.08 | 26.00 |
| **ITP** |  |  |  |  |  |  |  |  |  |  |
| ***Conditional Poisson Regression*** |  |  |  |  |  |  |  |  |  |  |
| *Cases in Risk Window* | 472 | | | | | 318 | | | | |
| *Cases in Control Window* | 196 | | | | | 99 | | | | |
| *Person-Days in Risk Window* | 40,277 | | | | | 27,697 | | | | |
| *Person-Days in Control Window* | 18,212 | | | | | 9,212 | | | | |
| *Model Results* | IRR | 95% CI | | P-Value | SE | IRR | 95% CI | | P-Value | SE |
|  | 1.12 | 0.94 | 1.33 | 0.21 | 0.09 | 1.08 | 0.86 | 1.37 | 0.50 | 0.12 |
| ***Attributable Risk*** |  |  |  |  |  |  |  |  |  |  |
| *# of Eligible Vaccinated* | 1,963,760 | | | | | 1,397,221 | | | | |
| *# of Vaccines* | 3,717,530 | | | | | 2,671,012 | | | | |
| *Person-Years* | 328,874 | | | | | 259,993 | | | | |
| *Attributable Risk per 100,000 Doses* | AR | 95% CI | | P-Value | SE | AR | 95% CI | | P-Value | SE |
|  | 1.33 | -0.73 | 3.38 | 0.21 | 1.05 | 0.92 | -1.73 | 3.56 | 0.50 | 1.35 |
| *Attributable Risk per 100,000 Person-Years* | AR | 95% CI | | P-Value | SE | AR | 95% CI | | P-Value | SE |
|  | 14.99 | -8.24 | 38.23 | 0.21 | 11.85 | 9.42 | -17.76 | 36.61 | 0.50 | 13.87 |
| **DIC** |  |  |  |  |  |  |  |  |  |  |
| ***Conditional Poisson Regression*** |  |  |  |  |  |  |  |  |  |  |
| *Cases in Risk Window* | 110 | | | | | 52 | | | | |
| *Cases in Control Window* | 65 | | | | | 27 | | | | |
| *Person-Days in Risk Window* | 6,555 | | | | | 3,082 | | | | |
| *Person-Days in Control Window* | 3,726 | | | | | 1,483 | | | | |
| *Model Results* | IRR | 95% CI | | P-Value | SE | IRR | 95% CI | | P-Value | SE |
|  | 1.36 | 0.90 | 2.06 | 0.15 | 0.21 | 1.79 | 0.97 | 3.29 | 0.06 | 0.31 |
| ***Attributable Risk*** |  |  |  |  |  |  |  |  |  |  |
| *# of Eligible Vaccinated* | 1,515,699 | | | | | 950,840 | | | | |
| *# of Vaccines* | 2,892,202 | | | | | 1,823,330 | | | | |
| *Person-Years* | 196,953 | | | | | 138,694 | | | | |
| *Attributable Risk per 100,000 Doses* | AR | 95% CI | | P-Value | SE | AR | 95% CI | | P-Value | SE |
|  | 1.00 | -0.32 | 2.33 | 0.14 | 0.68 | 1.26 | 0.08 | 2.44 | 0.04 | 0.60 |
| *Attributable Risk per 100,000 Person-Years* | AR | 95% CI | | P-Value | SE | AR | 95% CI | | P-Value | SE |
|  | 14.73 | -4.75 | 34.21 | 0.14 | 9.94 | 16.52 | 1.004 | 32.03 | 0.04 | 7.91 |

Abbreviations: IRR, incidence rate ratio; CI, confidence interval; SE, standard error; AR, attributable risk

**eTable 6. Summary of incidence rate ratio and attributable risk of AMI, inpatient PE, ITP, and DIC in primary SCCS analysis for the primary series COVID-19 vaccinations with post-vaccine control interval, adjusting for event-dependent observation time, seasonal variation in adverse event risk, outcome misclassification using MRR-derived PPV, and excluding cases with prior COVID-19**

|  | **BNT162b2** | | | | | **mRNA-1273** | | | | |
| --- | --- | --- | --- | --- | --- | --- | --- | --- | --- | --- |
| **AMI** |  |  |  |  |  |  |  |  |  |  |
| ***Conditional Poisson Regression*** |  |  |  |  |  |  |  |  |  |  |
| *Cases in Risk Window** | 787 | | | | | 302 | | | | |
| *Cases in Control Window** | 628 | | | | | 203 | | | | |
| *Person-Days in Risk Window* | 63,507 | | | | | 24,722 | | | | |
| *Person-Days in Control Window* | 47,126 | | | | | 14,828 | | | | |
| *Model Results* | IRR | 95% CI | | P-Value | SE | IRR | 95% CI | | P-Value | SE |
|  | 1.04 | 0.91 | 1.18 | 0.57 | 0.06 | 1.01 | 0.82 | 1.26 | 0.90 | 0.11 |
| ***Attributable Risk*** |  |  |  |  |  |  |  |  |  |  |
| *# of Eligible Vaccinated* | 442,459 | | | | | 179,969 | | | | |
| *# of Vaccines* | 842,181 | | | | | 342,257 | | | | |
| *Person-Years* | 56,803 | | | | | 25,921 | | | | |
| *Attributable Risk per 100,000 Doses* | AR | 95% CI | | P-Value | SE | AR | 95% CI | | P-Value | SE |
|  | 3.33 | -8.80 | 14.02 | 0.57 | 5.82 | 1.26 | -19.84 | 18.25 | 0.90 | 9.72 |
| *Attributable Risk per 100,000 Person-Years* | AR | 95% CI | | P-Value | SE | AR | 95% CI | | P-Value | SE |
|  | 49.34 | -130.53 | 207.87 | 0.57 | 86.33 | 16.68 | -261.94 | 240.96 | 0.90 | 128.29 |
| **PE (IP)** |  |  |  |  |  |  |  |  |  |  |
| ***Conditional Poisson Regression*** |  |  |  |  |  |  |  |  |  |  |
| *Cases in Risk Window** | 549 | | | | | 316 | | | | |
| *Cases in Control Window** | 403 | | | | | 201 | | | | |
| *Person-Days in Risk Window* | 43,617 | | | | | 26,266 | | | | |
| *Person-Days in Control Window* | 34,837 | | | | | 17,074 | | | | |
| *Model Results* | IRR | 95% CI | | P-Value | SE | IRR | 95% CI | | P-Value | SE |
|  | 1.19 | 1.03 | 1.38 | 0.02 | 0.08 | 1.15 | 0.94 | 1.41 | 0.17 | 0.10 |
| ***Attributable Risk*** |  |  |  |  |  |  |  |  |  |  |
| *# of Eligible Vaccinated* | 1,305,470 | | | | | 851,422 | | | | |
| *# of Vaccines* | 2,485,166 | | | | | 1,625,596 | | | | |
| *Person-Years* | 169,450 | | | | | 123,712 | | | | |
| *Attributable Risk per 100,000 Doses* | AR | 95% CI | | P-Value | SE | AR | 95% CI | | P-Value | SE |
|  | 3.50 | 0.55 | 6.05 | 0.01 | 1.40 | 2.58 | -1.24 | 5.69 | 0.14 | 1.77 |
| *Attributable Risk per 100,000 Person-Years* | AR | 95% CI | | P-Value | SE | AR | 95% CI | | P-Value | SE |
|  | 51.34 | 8.01 | 88.71 | 0.01 | 20.59 | 33.91 | -16.25 | 74.81 | 0.14 | 23.23 |
| **ITP** |  |  |  |  |  |  |  |  |  |  |
| ***Conditional Poisson Regression*** |  |  |  |  |  |  |  |  |  |  |
| *Cases in Risk Window* | ** | | | | | ** | | | | |
| *Cases in Control Window* | ** | | | | | ** | | | | |
| *Person-Days in Risk Window* | 1,460 | | | | | 1,112 | | | | |
| *Person-Days in Control Window* | 650 | | | | | 394 | | | | |
| *Model Results* | IRR | 95% CI | | P-Value | SE | IRR | 95% CI | | P-Value | SE |
|  | 2.15 | 0.42 | 10.95 | 0.35 | 0.83 | 1.31 | 0.23 | 7.53 | 0.76 | 0.89 |
| ***Attributable Risk*** |  |  |  |  |  |  |  |  |  |  |
| *# of Eligible Vaccinated* | 1,786,155 | | | | | 1,320,067 | | | | |
| *# of Vaccines* | 3,384,620 | | | | | 2,525,699 | | | | |
| *Person-Years* | 299,783 | | | | | 245,979 | | | | |
| *Attributable Risk per 100,000 Doses* | AR | 95% CI | | P-Value | SE | AR | 95% CI | | P-Value | SE |
|  | 0.31 | -0.79 | 0.53 | 0.35 | 0.34 | 0.12 | -1.78 | 0.46 | 0.83 | 0.57 |
| *Attributable Risk per 100,000 Person-Years* | AR | 95% CI | | P-Value | SE | AR | 95% CI | | P-Value | SE |
|  | 3.51 | -8.89 | 5.95 | 0.35 | 3.79 | 1.27 | -18.28 | 4.67 | 0.83 | 5.86 |
| **DIC** |  |  |  |  |  |  |  |  |  |  |
| ***Conditional Poisson Regression*** |  |  |  |  |  |  |  |  |  |  |
| *Cases in Risk Window* | 31 | | | | | ** | | | | |
| *Cases in Control Window* | 19 | | | | | ** | | | | |
| *Person-Days in Risk Window* | 1,944 | | | | | 931 | | | | |
| *Person-Days in Control Window* | 1,024 | | | | | 403 | | | | |
| *Model Results* | IRR | 95% CI | | P-Value | SE | IRR | 95% CI | | P-Value | SE |
|  | 1.18 | 0.44 | 3.13 | 0.74 | 0.50 | 1.21 | 0.25 | 5.94 | 0.82 | 0.81 |
| ***Attributable Risk*** |  |  |  |  |  |  |  |  |  |  |
| *# of Eligible Vaccinated* | 1,359,328 | | | | | 888,093 | | | | |
| *# of Vaccines* | 2,596,136 | | | | | 1,704,226 | | | | |
| *Person-Years* | 177,022 | | | | | 129,688 | | | | |
| *Attributable Risk per 100,000 Doses* | AR | 95% CI | | P-Value | SE | AR | 95% CI | | P-Value | SE |
|  | 0.18 | -1.50 | 0.81 | 0.76 | 0.59 | 0.14 | -2.43 | 0.66 | 0.86 | 0.79 |
| *Attributable Risk per 100,000 Person-Years* | AR | 95% CI | | P-Value | SE | AR | 95% CI | | P-Value | SE |
|  | 2.64 | -21.98 | 11.91 | 0.76 | 8.65 | 1.79 | -31.89 | 8.65 | 0.86 | 10.34 |

Abbreviations: IRR, incidence rate ratio; CI, confidence interval; SE, standard error; AR, attributable risk

** Cell suppressed to protect patient confidentiality

**eTable 7. Summary of incidence rate ratios and attributable risk of AMI, inpatient PE, ITP, and DIC in secondary SCCS analysis for the primary series COVID-19 vaccinations with pre-vaccination control interval, adjusting for event-dependent observation time and excluding cases with prior COVID-19**

|  | **BNT162b2** | | | | | **mRNA-1273** | | | | |
| --- | --- | --- | --- | --- | --- | --- | --- | --- | --- | --- |
| **AMI** |  |  |  |  |  |  |  |  |  |  |
| ***Conditional Poisson Regression*** |  |  |  |  |  |  |  |  |  |  |
| *Cases in Risk Window* | 914 | | | | | 351 | | | | |
| *Cases in Control Window* | 1,614 | | | | | 471 | | | | |
| *Person-Days in Risk Window* | 112,817 | | | | | 40,150 | | | | |
| *Person-Days in Control Window* | 156,421 | | | | | 48,028 | | | | |
| *Model Results* | IRR | 95% CI | | P-Value | SE | IRR | 95% CI | | P-Value | SE |
|  | 0.75 | 0.69 | 0.81 | <0.01 | 0.04 | 0.84 | 0.73 | 0.98 | 0.02 | 0.07 |
| ***Attributable Risk*** |  |  |  |  |  |  |  |  |  |  |
| *# of Eligible Vaccinated* | 472,851 | | | | | 189,689 | | | | |
| *# of Vaccines* | 888,538 | | | | | 355,933 | | | | |
| *Person-Years* | 59,054 | | | | | 26,558 | | | | |
| *Attributable Risk per 100,000 Doses* | AR | 95% CI | | P-Value | SE | AR | 95% CI | | P-Value | SE |
|  | -34.68 | -44.52 | -24.84 | <0.01 | 5.02 | -18.41 | -34.33 | -2.49 | 0.02 | 8.12 |
| *Attributable Risk per 100,000 Person-Years* | AR | 95% CI | | P-Value | SE | AR | 95% CI | | P-Value | SE |
|  | -521.78 | -669.85 | -373.71 | <0.01 | 75.55 | -246.74 | -460.12 | -33.35 | 0.02 | 108.87 |
| **PE (IP)** |  |  |  |  |  |  |  |  |  |  |
| ***Conditional Poisson Regression*** |  |  |  |  |  |  |  |  |  |  |
| *Cases in Risk Window* | 596 | | | | | 345 | | | | |
| *Cases in Control Window* | 953 | | | | | 419 | | | | |
| *Person-Days in Risk Window* | 70,896 | | | | | 38,617 | | | | |
| *Person-Days in Control Window* | 100,850 | | | | | 46,769 | | | | |
| *Model Results* | IRR | 95% CI | | P-Value | SE | IRR | 95% CI | | P-Value | SE |
|  | 0.89 | 0.80 | 0.99 | 0.03 | 0.05 | 0.99 | 0.86 | 1.15 | 0.90 | 0.08 |
| ***Attributable Risk*** |  |  |  |  |  |  |  |  |  |  |
| *# of Eligible Vaccinated* | 1,378,158 | | | | | 886,751 | | | | |
| *# of Vaccines* | 2,582,922 | | | | | 1,667,174 | | | | |
| *Person-Years* | 173,060 | | | | | 124,751 | | | | |
| *Attributable Risk per 100,000 Doses* | AR | 95% CI | | P-Value | SE | AR | 95% CI | | P-Value | SE |
|  | -2.86 | -5.44 | -0.28 | 0.03 | 1.32 | -0.20 | -3.25 | 2.86 | 0.90 | 1.56 |
| *Attributable Risk per 100,000 Person-Years* | AR | 95% CI | | P-Value | SE | AR | 95% CI | | P-Value | SE |
|  | -42.68 | -81.17 | -4.18 | 0.03 | 19.64 | -2.62 | -43.39 | 38.16 | 0.90 | 20.80 |
| **ITP** |  |  |  |  |  |  |  |  |  |  |
| ***Conditional Poisson Regression*** |  |  |  |  |  |  |  |  |  |  |
| *Cases in Risk Window* | 407 | | | | | 284 | | | | |
| *Cases in Control Window* | 394 | | | | | 227 | | | | |
| *Person-Days in Risk Window* | 48,436 | | | | | 34,233 | | | | |
| *Person-Days in Control Window* | 55,236 | | | | | 32,608 | | | | |
| *Model Results* | IRR | 95% CI | | P-Value | SE | IRR | 95% CI | | P-Value | SE |
|  | 1.17 | 1.01 | 1.34 | 0.03 | 0.07 | 1.18 | 0.99 | 1.41 | 0.07 | 0.09 |
| ***Attributable Risk*** |  |  |  |  |  |  |  |  |  |  |
| *# of Eligible Vaccinated* | 1,891,141 | | | | | 1,377,082 | | | | |
| *# of Vaccines* | 3,526,487 | | | | | 2,593,132 | | | | |
| *Person-Years* | 306,658 | | | | | 248,197 | | | | |
| *Attributable Risk per 100,000 Doses* | AR | 95% CI | | P-Value | SE | AR | 95% CI | | P-Value | SE |
|  | 1.65 | 0.14 | 3.16 | 0.03 | 0.77 | 1.66 | -0.12 | 3.43 | 0.07 | 0.91 |
| *Attributable Risk per 100,000 Person-Years* | AR | 95% CI | | P-Value | SE | AR | 95% CI | | P-Value | SE |
|  | 18.96 | 1.60 | 36.32 | 0.03 | 8.86 | 17.30 | -1.26 | 35.86 | 0.07 | 9.47 |
| **DIC** |  |  |  |  |  |  |  |  |  |  |
| ***Conditional Poisson Regression*** |  |  |  |  |  |  |  |  |  |  |
| *Cases in Risk Window* | 68 | | | | | 30 | | | | |
| *Cases in Control Window* | 61 | | | | | 26 | | | | |
| *Person-Days in Risk Window* | 4,999 | | | | | 2,441 | | | | |
| *Person-Days in Control Window* | 6,292 | | | | | 2,737 | | | | |
| *Model Results* | IRR | 95% CI | | P-Value | SE | IRR | 95% CI | | P-Value | SE |
|  | 1.07 | 0.71 | 1.61 | 0.75 | 0.21 | 1.30 | 0.67 | 2.51 | 0.43 | 0.34 |
| ***Attributable Risk*** |  |  |  |  |  |  |  |  |  |  |
| *# of Eligible Vaccinated* | 1,434,612 | | | | | 925,544 | | | | |
| *# of Vaccines* | 2,696,964 | | | | | 1,748,458 | | | | |
| *Person-Years* | 180,693 | | | | | 130,811 | | | | |
| *Attributable Risk per 100,000 Doses* | AR | 95% CI | | P-Value | SE | AR | 95% CI | | P-Value | SE |
|  | 0.16 | -0.84 | 1.17 | 0.75 | 0.51 | 0.40 | -0.61 | 1.40 | 0.44 | 0.51 |
| *Attributable Risk per 100,000 Person-Years* | AR | 95% CI | | P-Value | SE | AR | 95% CI | | P-Value | SE |
|  | 2.40 | -12.58 | 17.39 | 0.75 | 7.65 | 5.32 | -8.14 | 18.78 | 0.44 | 6.87 |

Abbreviations: IRR, incidence rate ratio; CI, confidence interval; SE, standard error; AR, attributable risk

**eTable 8. Summary of incidence rate ratios and attributable risk of AMI, inpatient PE, and ITP in exploratory SCRI analysis for the primary series COVID-19 vaccinations with pre-vaccination control interval, adjusting for event-dependent observation time and outcome misclassification using MRR-derived PPV**

|  | **BNT162b2** | | | | | **mRNA-1273** | | | | |
| --- | --- | --- | --- | --- | --- | --- | --- | --- | --- | --- |
| **AMI** |  |  |  |  |  |  |  |  |  |  |
| ***Conditional Poisson Regression*** |  |  |  |  |  |  |  |  |  |  |
| *Cases in Risk Window* | 1,353 | | | | | 436 | | | | |
| *Cases in Control Window* | 782 | | | | | 207 | | | | |
| *Person-Days in Risk Window* | 88,266 | | | | | 29,558 | | | | |
| *Person-Days in Control Window* | 59,767 | | | | | 17,999 | | | | |
| *Model Results* | IRR | 95% CI | | P-Value | SE | IRR | 95% CI | | P-Value | SE |
|  | 0.87 | 0.78 | 0.96 | <0.01 | 0.05 | 1.02 | 0.85 | 1.24 | 0.82 | 0.10 |
| ***Attributable Risk*** |  |  |  |  |  |  |  |  |  |  |
| *# of Eligible Vaccinated* | 544,380 | | | | | 208,562 | | | | |
| *# of Vaccines* | 1,025,254 | | | | | 391,503 | | | | |
| *Person-Years* | 68,128 | | | | | 29,219 | | | | |
| *Attributable Risk per 100,000 Doses* | AR | 95% CI | | P-Value | SE | AR | 95% CI | | P-Value | SE |
|  | -20.50 | -36.27 | -6.21 | 0.01 | 7.67 | 2.47 | -20.18 | 21.22 | 0.82 | 10.56 |
| *Attributable Risk per 100,000 Person-Years* | AR | 95% CI | | P-Value | SE | AR | 95% CI | | P-Value | SE |
|  | -308.53 | -545.88 | -93.43 | 0.01 | 115.42 | 33.09 | -270.34 | 284.30 | 0.82 | 141.49 |
| **PE (IP)** |  |  |  |  |  |  |  |  |  |  |
| ***Conditional Poisson Regression*** |  |  |  |  |  |  |  |  |  |  |
| *Cases in Risk Window* | 875 | | | | | 431 | | | | |
| *Cases in Control Window* | 517 | | | | | 169 | | | | |
| *Person-Days in Risk Window* | 60,585 | | | | | 29,451 | | | | |
| *Person-Days in Control Window* | 38,964 | | | | | 16,809 | | | | |
| *Model Results* | IRR | 95% CI | | P-Value | SE | IRR | 95% CI | | P-Value | SE |
|  | 0.95 | 0.84 | 1.07 | 0.41 | 0.06 | 1.28 | 1.05 | 1.57 | 0.01 | 0.10 |
| ***Attributable Risk*** |  |  |  |  |  |  |  |  |  |  |
| *# of Eligible Vaccinated* | 1,492,705 | | | | | 931,639 | | | | |
| *# of Vaccines* | 2,797,464 | | | | | 1,751,614 | | | | |
| *Person-Years* | 187,380 | | | | | 131,061 | | | | |
| *Attributable Risk per 100,000 Doses* | AR | 95% CI | | P-Value | SE | AR | 95% CI | | P-Value | SE |
|  | -1.62 | -5.83 | 2.11 | 0.42 | 2.03 | 5.45 | 1.23 | 8.91 | 0.01 | 1.96 |
| *Attributable Risk per 100,000 Person-Years* | AR | 95% CI | | P-Value | SE | AR | 95% CI | | P-Value | SE |
|  | -24.25 | -87.07 | 31.45 | 0.42 | 30.24 | 72.84 | 16.41 | 119.10 | 0.01 | 26.20 |
| **ITP** |  |  |  |  |  |  |  |  |  |  |
| ***Conditional Poisson Regression*** |  |  |  |  |  |  |  |  |  |  |
| *Cases in Risk Window* | ** | | | | | ** | | | | |
| *Cases in Control Window* | ** | | | | | ** | | | | |
| *Person-Days in Risk Window* | 1,868 | | | | | 1,339 | | | | |
| *Person-Days in Control Window* | 1,300 | | | | | 857 | | | | |
| *Model Results* | IRR | 95% CI | | P-Value | SE | IRR | 95% CI | | P-Value | SE |
|  | 1.78 | 0.522 | 6.06 | 0.36 | 0.63 | 1.93 | 0.40 | 9.31 | 0.41 | 0.80 |
| ***Attributable Risk*** |  |  |  |  |  |  |  |  |  |  |
| *# of Eligible Vaccinated* | 2,008,156 | | | | | 1,427,663 | | | | |
| *# of Vaccines* | 3,743,819 | | | | | 2,687,855 | | | | |
| *Person-Years* | 325,466 | | | | | 257,205 | | | | |
| *Attributable Risk per 100,000 Doses* | AR | 95% CI | | P-Value | SE | AR | 95% CI | | P-Value | SE |
|  | 0.26 | -0.55 | 0.50 | 0.33 | 0.27 | 0.28 | -0.86 | 0.51 | 0.43 | 0.35 |
| *Attributable Risk per 100,000 Person-Years* | AR | 95% CI | | P-Value | SE | AR | 95% CI | | P-Value | SE |
|  | 3.00 | -6.27 | 5.72 | 0.33 | 3.06 | 2.88 | -8.95 | 5.34 | 0.43 | 3.64 |

Abbreviations: IRR, incidence rate ratio; CI, confidence interval; SE, standard error; AR, attributable risk

** Cell suppressed to protect patient confidentiality

**eTable 9. Summary of incidence rate ratios and attributable risk of AMI, PE, ITP, Myo-/Peri and BP in primary SCCS analysis with post-vaccine control interval, adjusting for event-dependent observation time for the booster dose COVID-19 vaccine**

| **Analysis** | **BNT162b2** | | | | | **mRNA-1273** | | | | |
| --- | --- | --- | --- | --- | --- | --- | --- | --- | --- | --- |
| ***AMI*** | | | | | | | | | | |
| ***Conditional Poisson Regression*** | | | | | | | | | | |
| *Cases in Risk Window* | 2,459 | | | | | 2,422 | | | | |
| *Cases in Control Window* | 5,642 | | | | | 5,519 | | | | |
| *Person-Days in Risk Window* | 223,937 | | | | | 219,792 | | | | |
| *Person-Days in Control Window* | 445,349 | | | | | 440,002 | | | | |
| *Model Results* | *IRR* | *95% CI* | | *P-Value* | *SE* | *IRR* | *95% CI* | | *P-Value* | *SE* |
|  | 1.002 | 0.95 | 1.05 | 0.94 | 0.03 | 1.01 | 0.96 | 1.07 | 0.62 | 0.03 |
| ***Attributable Risk*** | | | | | | | | | | |
| *# of Eligible Vaccinated* | 2,990,542 | | | | | 3,106,236 | | | | |
| *Person-Years* | 228,477 | | | | | 237,167 | | | | |
| *Attributable Risk per 100,000 Doses* | *AR* | *95% CI* | | *P-Value* | *SE* | *AR* | *95% CI* | | *P-Value* | *SE* |
|  | 0.15 | -3.97 | 4.26 | 0.94 | 2.10 | 0.98 | -2.90 | 4.26 | 0.62 | 1.98 |
| *Attributable Risk per 100,000 Person-Years* | *AR* | *95% CI* | |  | *SE* | *AR* | *95% CI* | |  | *SE* |
|  | 1.94 | -51.92 | 55.80 |  | 27.48 | 12.79 | -38.04 | -38.04 |  | 25.93 |
| ***ITP*** | | | | | | | | | | |
| ***Conditional Poisson Regression*** | | | | | | | | | | |
| *Cases in Risk Window* | 24 | | | | | 22 | | | | |
| *Cases in Control Window* | 25 | | | | | 17 | | | | |
| *Person-Days in Risk Window* | 2,058 | | | | | 1,638 | | | | |
| *Person-Days in Control Window* | 2,299 | | | | | 1,815 | | | | |
| *Model Results* | *IRR* | *95% CI* | | *P-Value* | *SE* | *IRR* | *95% CI* | | *P-Value* | *SE* |
|  | 1.17 | 0.66 | 2.04 | 0.59 | 0.29 | 1.54 | 0.82 | 2.91 | 0.18 | 0.32 |
| ***Attributable Risk*** | | | | | | | | | | |
| *# of Eligible Vaccinated* | 2,990,542 | | | | | 3,106,236 | | | | |
| *Person-Years* | 8,193 | | | | | 8,510 | | | | |
| *Attributable Risk per 100,000 Doses* | *AR* | *95% CI* | | *P-Value* | *SE* | *AR* | *95% CI* | | *P-Value* | *SE* |
|  | 0.11 | -0.30 | 0.53 | 0.59 | 0.21 | 0.25 | -0.11 | 0.61 | 0.18 | 0.18 |
| *Attributable Risk per 100,000 Person-Years* | *AR* | *95% CI* | |  | *SE* | *AR* | *95% CI* | |  | *SE* |
|  | 0.997 | -2.62 | 4.61 |  | 1.85 | 2.18 | -0.97 | 5.34 |  | 1.61 |
| ***PE*** | | | | | | | | | | |
| ***Conditional Poisson Regression*** | | | | | | | | | | |
| *Cases in Risk Window* | 724 | | | | | 657 | | | | |
| *Cases in Control Window* | 1,898 | | | | | 1,806 | | | | |
| *Person-Days in Risk Window* | 72,748 | | | | | 68,381 | | | | |
| *Person-Days in Control Window* | 146,463 | | | | | 138,425 | | | | |
| *Model Results* | *IRR* | *95% CI* | | *P-Value* | *SE* | *IRR* | *95% CI* | | *P-Value* | *SE* |
|  | 0.87 | 0.79 | 0.95 | <0.01 | 0.05 | 0.84 | 0.76 | 0.92 | <0.01 | 0.05 |
| ***Attributable Risk*** | | | | | | | | | | |
| *# of Eligible Vaccinated* | 2,990,542 | | | | | 3,106,236 | | | | |
| *Person-Years* | 228,477 | | | | | 237,167 | | | | |
| *Attributable Risk per 100,000 Doses* | *AR* | *95% CI* | | *P-Value* | *SE* | *AR* | *95% CI* | | *P-Value* | *SE* |
|  | -3.71 | -5.99 | -1.44 | <0.01 | 1.16 | -4.08 | -6.15 | -2.01 | <0.01 | 1.06 |
| *Attributable Risk per 100,000 Person-Years* | *AR* | *95% CI* | |  | *SE* | *AR* | *95% CI* | |  | *SE* |
|  | -48.62 | -78.44 | -18.79 |  | 15.22 | -53.47 | -80.59 | -26.36 |  | 13.83 |
| ***Myo-/Peri*** | | | | | | | | | | |
| ***Conditional Poisson Regression*** | | | | | | | | | | |
| *Cases in Risk Window* | 166 | | | | | 144 | | | | |
| *Cases in Control Window* | 517 | | | | | 468 | | | | |
| *Person-Days in Risk Window* | 14,309 | | | | | 12,844 | | | | |
| *Person-Days in Control Window* | 45,610 | | | | | 41,068 | | | | |
| *Model Results* | *IRR* | *95% CI* | | *P-Value* | *SE* | *IRR* | *95% CI* | | *P-Value* | *SE* |
|  | 1.09 | 0.91 | 1.30 | 0.35 | 0.09 | 1.07 | 0.89 | 1.30 | 0.45 | 0.10 |
| ***Attributable Risk*** | | | | | | | | | | |
| *# of Eligible Vaccinated* | 3,004,387 | | | | | 3,124,213 | | | | |
| *Person-Years* | 172,351 | | | | | 179,143 | | | | |
| *Attributable Risk per 100,000 Doses* | *AR* | *95% CI* | | *P-Value* | *SE* | *AR* | *95% CI* | | *P-Value* | *SE* |
|  | 0.45 | -0.51 | 1.42 | 0.36 | 0.49 | 0.32 | -0.52 | 1.16 | 0.45 | 0.43 |
| *Attributable Risk per 100,000 Person-Years* | *AR* | *95% CI* | |  | *SE* | *AR* | *95% CI* | |  | *SE* |
|  | 7.90 | -8.95 | 24.75 |  | 8.60 | 5.61 | -9.10 | 20.31 |  | 7.50 |
| ***BP*** | | | | | | | | | | |
| ***Conditional Poisson Regression*** | | | | | | | | | | |
| *Cases in Risk Window* | 827 | | | | | 752 | | | | |
| *Cases in Control Window* | 847 | | | | | 842 | | | | |
| *Person-Days in Risk Window* | 70,198 | | | | | 66,890 | | | | |
| *Person-Days in Control Window* | 78,726 | | | | | 75,236 | | | | |
| *Model Results* | *IRR* | *95% CI* | | *P-Value* | *SE* | *IRR* | *95% CI* | | *P-Value* | *SE* |
|  | 1.13 | 1.03 | 1.25 | 0.01 | 0.05 | 1.03 | 0.93 | 1.13 | 0.60 | 0.05 |
| ***Attributable Risk*** | | | | | | | | | | |
| *# of Eligible Vaccinated* | 3,016,489 | | | | | 3,139,611 | | | | |
| *Person-Years* | 344,867 | | | | | 358,514 | | | | |
| *Attributable Risk per 100,000 Doses* | *AR* | *95% CI* | | *P-Value* | *SE* | *AR* | *95% CI* | | *P-Value* | *SE* |
|  | 3.21 | 0.74 | 5.69 | 0.01 | 1.26 | 0.63 | -1.72 | 2.98 | 0.60 | 1.20 |
| *Attributable Risk per 100,000 Person-Years* | *AR* | *95% CI* | |  | *SE* | *AR* | *95% CI* | |  | *SE* |
|  | 28.10 | 6.47 | 49.73 |  | 11.03 | 5.48 | -15.10 | 26.07 |  | 10.50 |

Abbreviations: IRR, incidence rate ratio; CI, confidence interval; SE, standard error; AR, attributable risk

**eTable 10. Summary of incidence rate ratios and attributable risk of AMI, PE, ITP, Myo-/Peri and BP in primary SCCS analysis with post-vaccine control interval, adjusting for event-dependent observation time, seasonal variation in adverse event risk, and excluding cases with prior COVID-19 for the booster dose COVID-19 vaccine**

| **Analysis** | **BNT162b2** | | | | | **mRNA-1273** | | | | |
| --- | --- | --- | --- | --- | --- | --- | --- | --- | --- | --- |
| ***AMI*** | | | | | | | | | | |
| ***Conditional Poisson Regression*** | | | | | | | | | | |
| *Cases in Risk Window* | 2,138 | | | | | 2,129 | | | | |
| *Cases in Control Window* | 4,880 | | | | | 4,871 | | | | |
| *Person-Days in Risk Window* | 194,165 | | | | | 193,843 | | | | |
| *Person-Days in Control Window* | 388,690 | | | | | 389,858 | | | | |
| *Model Results* | *IRR* | *95% CI* | | *P-Value* | *SE* | *IRR* | *95% CI* | | *P-Value* | *SE* |
|  | 1.06 | 1.003 | 1.12 | 0.04 | 0.03 | 1.05 | 0.998 | 1.11 | 0.06 | 0.03 |
| ***Attributable Risk*** | | | | | | | | | | |
| *# of Eligible Vaccinated* | 2,990,542 | | | | | 3,106,236 | | | | |
| *Person-Years* | 228,477 | | | | | 237,167 | | | | |
| *Attributable Risk per 100,000 Doses* | *AR* | *95% CI* | | *P-Value* | *SE* | *AR* | *95% CI* | | *P-Value* | *SE* |
|  | 3.91 | 0.19 | 7.63 | 0.04 | 1.90 | 3.42 | -0.17 | 7.63 | 0.06 | 1.84 |
| *Attributable Risk per 100,000 Person-Years* | *AR* | *95% CI* | |  | *SE* | *AR* | *95% CI* | |  | *SE* |
|  | 51.18 | 2.47 | 99.89 |  | 24.85 | 44.84 | -2.28 | -2.28 |  | 24.04 |
| ***ITP*** | | | | | | | | | | |
| ***Conditional Poisson Regression*** | | | | | | | | | | |
| *Cases in Risk Window* | 20 | | | | | 20 | | | | |
| *Cases in Control Window* | 23 | | | | | 17 | | | | |
| *Person-Days in Risk Window* | 1,806 | | | | | 1,554 | | | | |
| *Person-Days in Control Window* | 2,011 | | | | | 1,719 | | | | |
| *Model Results* | *IRR* | *95% CI* | | *P-Value* | *SE* | *IRR* | *95% CI* | | *P-Value* | *SE* |
|  | 1.13 | 0.62 | 2.07 | 0.69 | 0.31 | 1.50 | 0.78 | 2.87 | 0.22 | 0.33 |
| ***Attributable Risk*** | | | | | | | | | | |
| *# of Eligible Vaccinated* | 2,990,542 | | | | | 3,106,236 | | | | |
| *Person-Years* | 8,193 | | | | | 8,510 | | | | |
| *Attributable Risk per 100,000 Doses* | *AR* | *95% CI* | | *P-Value* | *SE* | *AR* | *95% CI* | | *P-Value* | *SE* |
|  | 0.08 | -0.29 | 0.45 | 0.68 | 0.19 | 0.21 | -0.14 | 0.56 | 0.23 | 0.18 |
| *Attributable Risk per 100,000 Person-Years* | *AR* | *95% CI* | |  | *SE* | *AR* | *95% CI* | |  | *SE* |
|  | 0.68 | -2.56 | 3.93 |  | 1.66 | 1.87 | -1.19 | 4.93 |  | 1.56 |
| ***PE*** | | | | | | | | | | |
| ***Conditional Poisson Regression*** | | | | | | | | | | |
| *Cases in Risk Window* | 601 | | | | | 569 | | | | |
| *Cases in Control Window* | 1,604 | | | | | 1,539 | | | | |
| *Person-Days in Risk Window* | 61,251 | | | | | 58,530 | | | | |
| *Person-Days in Control Window* | 124,202 | | | | | 118,694 | | | | |
| *Model Results* | *IRR* | *95% CI* | | *P-Value* | *SE* | *IRR* | *95% CI* | | *P-Value* | *SE* |
|  | 0.86 | 0.78 | 0.95 | <0.01 | 0.05 | 0.87 | 0.79 | 0.96 | 0.01 | 0.05 |
| ***Attributable Risk*** | | | | | | | | | | |
| *# of Eligible Vaccinated* | 2,990,542 | | | | | 3,106,236 | | | | |
| *Person-Years* | 228,477 | | | | | 237,167 | | | | |
| *Attributable Risk per 100,000 Doses* | *AR* | *95% CI* | | *P-Value* | *SE* | *AR* | *95% CI* | | *P-Value* | *SE* |
|  | -3.34 | -5.40 | -1.29 | <0.01 | 1.05 | -2.71 | -4.61 | -0.81 | 0.01 | 0.97 |
| *Attributable Risk per 100,000 Person-Years* | *AR* | *95% CI* | |  | *SE* | *AR* | *95% CI* | |  | *SE* |
|  | -43.77 | -70.64 | -16.91 |  | 13.71 | -35.50 | -60.41 | -10.59 |  | 12.71 |
| ***Myo-/Peri*** | | | | | | | | | | |
| ***Conditional Poisson Regression*** | | | | | | | | | | |
| *Cases in Risk Window* | 148 | | | | | 132 | | | | |
| *Cases in Control Window* | 438 | | | | | 420 | | | | |
| *Person-Days in Risk Window* | 12,272 | | | | | 11,587 | | | | |
| *Person-Days in Control Window* | 39,182 | | | | | 37,203 | | | | |
| *Model Results* | *IRR* | *95% CI* | | *P-Value* | *SE* | *IRR* | *95% CI* | | *P-Value* | *SE* |
|  | 1.13 | 0.93 | 1.37 | 0.21 | 0.10 | 1.13 | 0.92 | 1.37 | 0.24 | 0.10 |
| ***Attributable Risk*** | | | | | | | | | | |
| *# of Eligible Vaccinated* | 3,004,387 | | | | | 3,124,213 | | | | |
| *Person-Years* | 172,351 | | | | | 179,143 | | | | |
| *Attributable Risk per 100,000 Doses* | *AR* | *95% CI* | | *P-Value* | *SE* | *AR* | *95% CI* | | *P-Value* | *SE* |
|  | 0.57 | -0.34 | 1.48 | 0.22 | 0.46 | 0.47 | -0.33 | 1.27 | 0.25 | 0.41 |
| *Attributable Risk per 100,000 Person-Years* | *AR* | *95% CI* | |  | *SE* | *AR* | *95% CI* | |  | *SE* |
|  | 9.90 | -5.93 | 25.74 |  | 8.08 | 8.23 | -5.75 | 22.22 |  | 7.14 |
| ***BP*** | | | | | | | | | | |
| ***Conditional Poisson Regression*** | | | | | | | | | | |
| *Cases in Risk Window* | 771 | | | | | 704 | | | | |
| *Cases in Control Window* | 787 | | | | | 787 | | | | |
| *Person-Days in Risk Window* | 65,330 | | | | | 62,581 | | | | |
| *Person-Days in Control Window* | 73,398 | | | | | 70,410 | | | | |
| *Model Results* | *IRR* | *95% CI* | | *P-Value* | *SE* | *IRR* | *95% CI* | | *P-Value* | *SE* |
|  | 1.17 | 1.06 | 1.29 | <0.01 | 0.05 | 1.16 | 1.05 | 1.29 | <0.01 | 0.05 |
| ***Attributable Risk*** | | | | | | | | | | |
| *# of Eligible Vaccinated* | 3,016,489 | | | | | 3,139,611 | | | | |
| *Person-Years* | 344,867 | | | | | 358,514 | | | | |
| *Attributable Risk per 100,000 Doses* | *AR* | *95% CI* | | *P-Value* | *SE* | *AR* | *95% CI* | | *P-Value* | *SE* |
|  | 3.73 | 1.38 | 6.08 | <0.01 | 1.20 | 3.16 | 1.02 | 5.30 | <0.01 | 1.09 |
| *Attributable Risk per 100,000 Person-Years* | *AR* | *95% CI* | |  | *SE* | *AR* | *95% CI* | |  | *SE* |
|  | 32.64 | 12.06 | 53.21 |  | 10.50 | 27.67 | 8.91 | 46.43 |  | 9.57 |

Abbreviations: IRR, incidence rate ratio; CI, confidence interval; SE, standard error; AR, attributable risk

**eTable 11. Summary of incidence rate ratios and attributable risk of AMI, PE, ITP, Myo-/Peri and BP in secondary SCCS analysis with pre- and post-vaccine control interval, adjusting for event-dependent observation time, seasonal variation in adverse event risk, and excluding cases with prior COVID-19 for the booster dose COVID-19 vaccine**

| **Analysis** | **BNT162b2** | | | | | **mRNA-1273** | | | | |
| --- | --- | --- | --- | --- | --- | --- | --- | --- | --- | --- |
| ***AMI*** | | | | | | | | | | |
| ***Conditional Poisson Regression*** | | | | | | | | | | |
| *Cases in Risk Window* | 2,129 | | | | | 2,123 | | | | |
| *Cases in Control Window* | 6,363 | | | | | 6,364 | | | | |
| *Person-Days in Risk Window* | 234,999 | | | | | 235,148 | | | | |
| *Person-Days in Control Window* | 713,176 | | | | | 715,886 | | | | |
| *Model Results* | *IRR* | *95% CI* | | *P-Value* | *SE* | *IRR* | *95% CI* | | *P-Value* | *SE* |
|  | 1.09 | 1.04 | 1.15 | <0.01 | 0.03 | 1.09 | 1.03 | 1.14 | <0.01 | 0.03 |
| ***Attributable Risk*** | | | | | | | | | | |
| *# of Eligible Vaccinated* | 2,966,500 | | | | | 3,082,590 | | | | |
| *Person-Years* | 226,619 | | | | | 235,336 | | | | |
| *Attributable Risk per 100,000 Doses* | *AR* | *95% CI* | | *P-Value* | *SE* | *AR* | *95% CI* | | *P-Value* | *SE* |
|  | 6.11 | 2.60 | 9.62 | <0.01 | 1.79 | 5.45 | 2.09 | 9.62 | <0.01 | 1.71 |
| *Attributable Risk per 100,000 Person-Years* | *AR* | *95% CI* | |  | *SE* | *AR* | *95% CI* | |  | *SE* |
|  | 80.02 | 34.09 | 125.96 |  | 23.44 | 71.42 | 27.43 | 27.43 |  | 22.45 |
| ***ITP*** | | | | | | | | | | |
| ***Conditional Poisson Regression*** | | | | | | | | | | |
| *Cases in Risk Window* | 19 | | | | | 19 | | | | |
| *Cases in Control Window* | 38 | | | | | 27 | | | | |
| *Person-Days in Risk Window* | 2,394 | | | | | 1,895 | | | | |
| *Person-Days in Control Window* | 5,077 | | | | | 4,035 | | | | |
| *Model Results* | *IRR* | *95% CI* | | *P-Value* | *SE* | *IRR* | *95% CI* | | *P-Value* | *SE* |
|  | 1.11 | 0.64 | 1.92 | 0.72 | 0.28 | 1.57 | 0.87 | 2.83 | 0.13 | 0.30 |
| ***Attributable Risk*** | | | | | | | | | | |
| *# of Eligible Vaccinated* | 2,959,608 | | | | | 3,074,932 | | | | |
| *Person-Years* | 8,127 | | | | | 8,445 | | | | |
| *Attributable Risk per 100,000 Doses* | *AR* | *95% CI* | | *P-Value* | *SE* | *AR* | *95% CI* | | *P-Value* | *SE* |
|  | 0.06 | -0.27 | 0.40 | 0.72 | 0.17 | 0.22 | -0.08 | 0.53 | 0.15 | 0.16 |
| *Attributable Risk per 100,000 Person-Years* | *AR* | *95% CI* | |  | *SE* | *AR* | *95% CI* | |  | *SE* |
|  | 0.54 | -2.39 | 3.48 |  | 1.50 | 1.96 | -0.73 | 4.66 |  | 1.38 |
| ***PE*** | | | | | | | | | | |
| ***Conditional Poisson Regression*** | | | | | | | | | | |
| *Cases in Risk Window* | 598 | | | | | 569 | | | | |
| *Cases in Control Window* | 2,062 | | | | | 1,968 | | | | |
| *Person-Days in Risk Window* | 73,947 | | | | | 70,424 | | | | |
| *Person-Days in Control Window* | 225,393 | | | | | 214,931 | | | | |
| *Model Results* | *IRR* | *95% CI* | | *P-Value* | *SE* | *IRR* | *95% CI* | | *P-Value* | *SE* |
|  | 0.93 | 0.84 | 1.02 | 0.11 | 0.05 | 0.95 | 0.87 | 1.05 | 0.34 | 0.05 |
| ***Attributable Risk*** | | | | | | | | | | |
| *# of Eligible Vaccinated* | 2,966,500 | | | | | 3,082,590 | | | | |
| *Person-Years* | 226,619 | | | | | 235,336 | | | | |
| *Attributable Risk per 100,000 Doses* | *AR* | *95% CI* | | *P-Value* | *SE* | *AR* | *95% CI* | | *P-Value* | *SE* |
|  | -1.60 | -3.48 | 0.27 | 0.09 | 0.96 | -0.87 | -2.61 | 0.87 | 0.33 | 0.89 |
| *Attributable Risk per 100,000 Person-Years* | *AR* | *95% CI* | |  | *SE* | *AR* | *95% CI* | |  | *SE* |
|  | -21.00 | -45.59 | 3.59 |  | 12.55 | -11.41 | -34.23 | 11.40 |  | 11.64 |
| ***Myo-/Peri*** | | | | | | | | | | |
| ***Conditional Poisson Regression*** | | | | | | | | | | |
| *Cases in Risk Window* | 148 | | | | | 132 | | | | |
| *Cases in Control Window* | 577 | | | | | 552 | | | | |
| *Person-Days in Risk Window* | 15,186 | | | | | 14,359 | | | | |
| *Person-Days in Control Window* | 63,643 | | | | | 60,421 | | | | |
| *Model Results* | *IRR* | *95% CI* | | *P-Value* | *SE* | *IRR* | *95% CI* | | *P-Value* | *SE* |
|  | 1.10 | 0.92 | 1.32 | 0.31 | 0.09 | 1.08 | 0.90 | 1.31 | 0.40 | 0.10 |
| ***Attributable Risk*** | | | | | | | | | | |
| *# of Eligible Vaccinated* | 2,984,166 | | | | | 3,103,944 | | | | |
| *Person-Years* | 171,174 | | | | | 177,959 | | | | |
| *Attributable Risk per 100,000 Doses* | *AR* | *95% CI* | | *P-Value* | *SE* | *AR* | *95% CI* | | *P-Value* | *SE* |
|  | 0.45 | -0.44 | 1.33 | 0.32 | 0.45 | 0.33 | -0.46 | 1.13 | 0.41 | 0.41 |
| *Attributable Risk per 100,000 Person-Years* | *AR* | *95% CI* | |  | *SE* | *AR* | *95% CI* | |  | *SE* |
|  | 7.78 | -7.66 | 23.23 |  | 7.88 | 5.79 | -8.08 | 19.66 |  | 7.08 |
| ***BP*** | | | | | | | | | | |
| ***Conditional Poisson Regression*** | | | | | | | | | | |
| *Cases in Risk Window* | 767 | | | | | 699 | | | | |
| *Cases in Control Window* | 1,487 | | | | | 1,558 | | | | |
| *Person-Days in Risk Window* | 94,526 | | | | | 94,701 | | | | |
| *Person-Days in Control Window* | 201,106 | | | | | 201,347 | | | | |
| *Model Results* | *IRR* | *95% CI* | | *P-Value* | *SE* | *IRR* | *95% CI* | | *P-Value* | *SE* |
|  | 1.13 | 1.03 | 1.23 | 0.01 | 0.04 | 1.04 | 0.95 | 1.13 | 0.44 | 0.05 |
| ***Attributable Risk*** | | | | | | | | | | |
| *# of Eligible Vaccinated* | 2,985,124 | | | | | 3,107,759 | | | | |
| *Person-Years* | 341,256 | | | | | 354,846 | | | | |
| *Attributable Risk per 100,000 Doses* | *AR* | *95% CI* | | *P-Value* | *SE* | *AR* | *95% CI* | | *P-Value* | *SE* |
|  | 2.88 | 0.71 | 5.05 | 0.01 | 1.11 | 0.78 | -1.24 | 2.79 | 0.45 | 1.03 |
| *Attributable Risk per 100,000 Person-Years* | *AR* | *95% CI* | |  | *SE* | *AR* | *95% CI* | |  | *SE* |
|  | 25.17 | 6.19 | 44.15 |  | 9.68 | 6.81 | -10.83 | 24.45 |  | 9.00 |

Abbreviations: IRR, incidence rate ratio; CI, confidence interval; SE, standard error; AR, attributable risk
